# Partial dependence of ultrasonically estimated fetal weight on biometric parameters

**DOI:** 10.1101/2024.09.17.24313697

**Authors:** Vasiliki Bitsouni, Nikolaos Gialelis, Vasilis Tsilidis

## Abstract

**Objectives:** To assess the contribution of each sonographically measured parameter on fetal weight formulas.

**Methods:** Multiple datasets and datasets extracted from existing literature are employed in this study. The Sobol’ method is then implemented on each of these datasets. Following this, bootstrapping is carried out using the indices from individual weeks as sample points. The mean value for each resulting sample is calculated. Finally, the median, along with a 95% confidence interval, is estimated based on the empirical distribution. This methodology is repeated for all formulas examined.

**Results:** The median, along with a 95% CI, for the estimate of the mean of the Sobol’ sensitivity indices for the parameters of 29 known formulas are presented.

**Conclusions:** Depending on the formula, the values of some parameters show substantial fluctuations or are insignificant to the fetal weight estimation.

## 1 Introduction

Estimating the weight of a fetus is a critical aspect of prenatal care, primarily conducted through ultrasound imaging. This method allows healthcare providers to measure various fetal parameters, such as biparietal diameter (BPD), abdominal circumference (AC), head circumference (HC), and femur length (FL). These measurements are then used as parameters in established mathematical formulas to calculate the estimated fetal weight (EFW) (see Figure 1).

**Figure 1:**
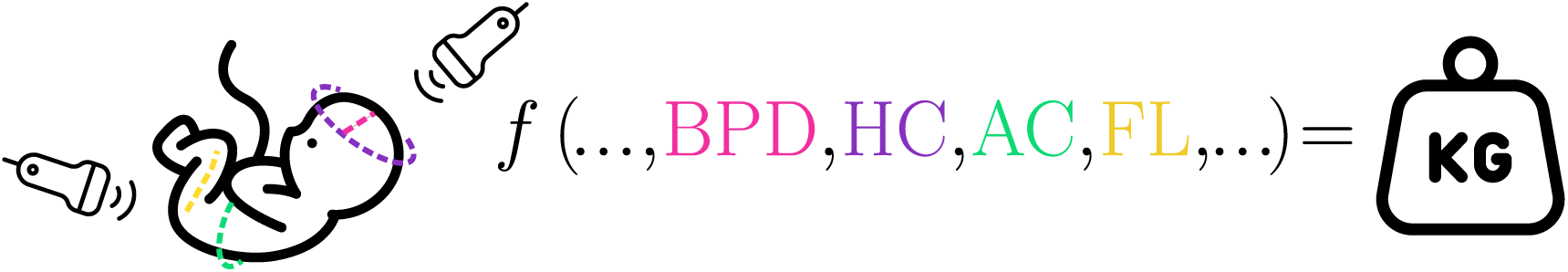
The biometric characteristics of a fetus, such as the circumference of its head and abdomen, the length of its femur, or its biparietal diameter are measured using ultrasound. The measurements are then entered into various mathematical formulas to estimate the weight of the fetus.

The parameters of a mathematical function do not necessarily contribute equally to its output. In many mathematical models, particularly nonlinear ones, certain parameters can have a more significant impact on the result than others. Statistical techniques can reveal the varying degrees of influence each parameter has, illustrating that the overall result is often a product of both individual and interactive effects among the parameters involved. This complexity underscores the importance of analyzing parameter contributions rather than assuming uniform influence across all inputs.

There exist many studies comparing the accuracy of different formulas for the calculation of EFW^[22;40;23;5;18;15;39;48;53]^. They often involve retrospective and prospective analysis, comparing predicted weights to actual birth weights to assess accuracy, bias, and reproducibility. Such articles underscore the ongoing quest for more reliable fetal weight estimation techniques that can enhance obstetric care by improving predictions related to delivery management and neonatal outcomes. However, a study that systematically assess the contribution of the involved parameters of each formula to the EFW does not exist.

The scope of the present study is to evaluate the effect of the biometric parameters of various formulas on the EFW. The utilized approach relies on an unbiased global sensitivity analysis scheme, known as the Sobol’ method.

## 2 Methods

An overview of the methods employed in the present study can be found in Figure 2. Subsequently, a detailed description of the methods is presented.

**Figure 2:**
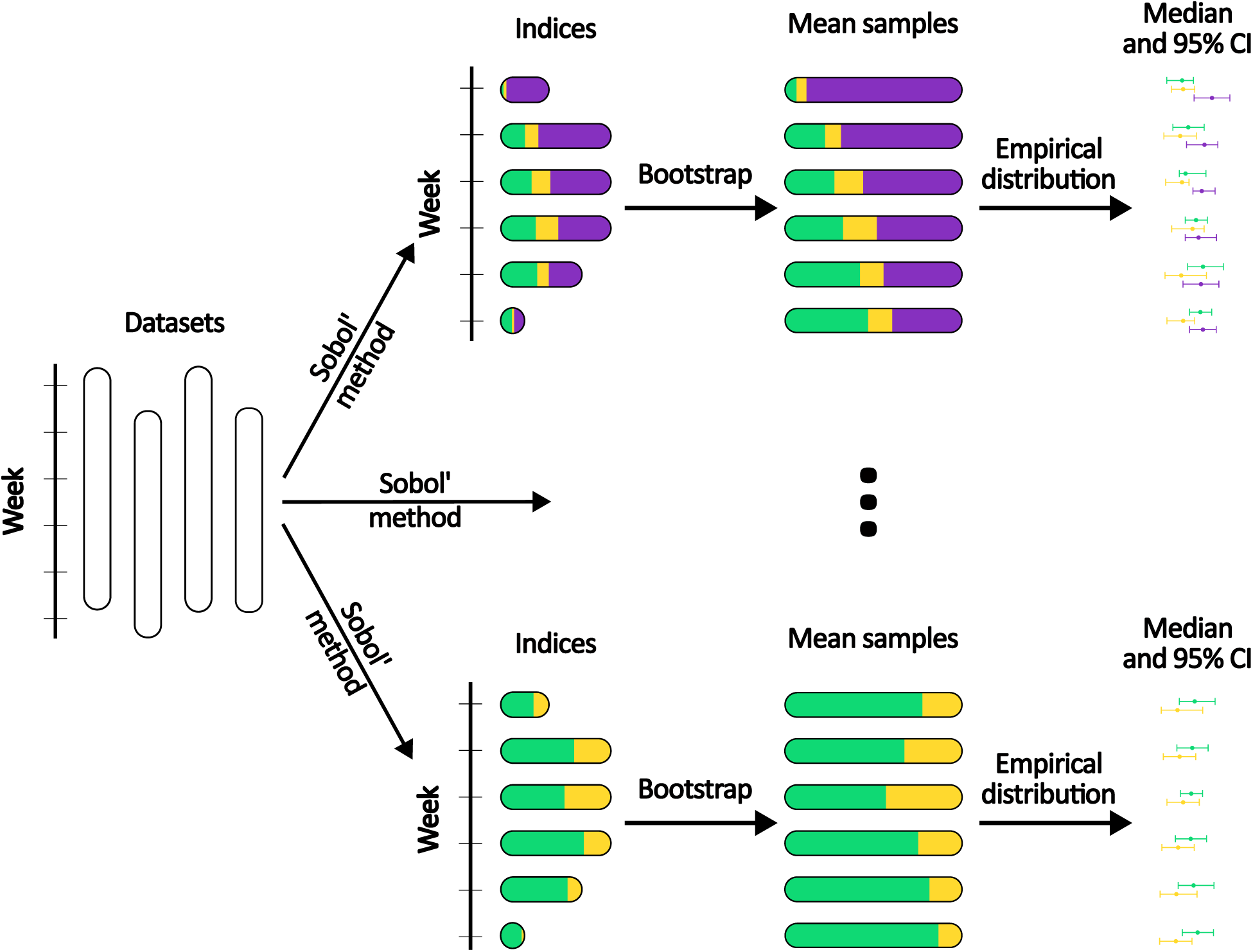
The current study employs a multi-step process involving the utilization of numerous datasets sourced from existing literature. The Sobol’ method is then applied to each of these datasets, for all the studied formulas. Following this, bootstrapping is conducted using the indices from each individual week as the sample points. The mean value is then calculated for each of the resulting samples. Finally, the median, along with a 95% confidence interval, are estimated using the empirical distribution.

### 2.1 Sensitivity analysis

The Sobol’ method, a global sensitivity analysis technique, is a variance-based approach employed to evaluate how changes in parameter inputs influence the outputs of a mathematical model. The method operates by partitioning the variance of the outputs generated by the model into distinct contributions from individual input parameters and their respective interactions. Assuming that *f* is the mathematical formula used for fetal weight estimation and **X** is the vector of the parameters of *f*, then by the Hoeffding decomposition, the total variance of the formula can be written as:

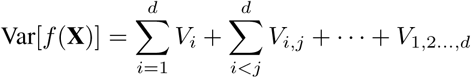

where *V_i_* represents the variance due to the main effect of parameter *i*, *V_ij_* represents the variance due to the interaction between parameters *i* and *j*, and so on.

Sobol’ sensitivity indices are normalized metrics that correspond to the aforementioned representations. The first-order sensitivity indices are defined as

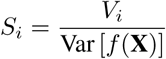

A high first-order index indicates that the parameter has a significant direct influence on the output. The second-order sensitivity indices are defined as

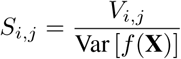

A high second-order index indicates that the combined effects of the two parameters are not merely additive. Both the first- and second-order indices range from 0 to 1.

The Julia package GlobalSensitivity.jl^[12]^ was employed for the calculation of the Sobol’ indices.

### 2.2 Sampling

In order to eliminate bias from the dataset and at the same time investigate the whole parameter space uniformly (i.e., more evenly), low-discrepancy synthetic data were generated. A quasi-random low-discrepancy sequence of numbers was generated in the interval between the 10th and the 90th percentile of each parameter, utilizing the Sobol’ sampling method, which is described in Sobol^[49]^. The Julia package QuasiMonteCarlo.jl was employed to generate a Sobol’ sequence of 10^6^ numbers for each parameter.

### 2.3 Data

A rundown of the utilized datasets can be seen in Table 1. In all the studies presented in Table 1, a number of measurements, regarding the parameters of interest, were taken from pregnant women of various ethnicities at various gestational ages. Furthermore, 10th and 90th percentile charts were calculated for all parameters. In the present study, the 10th and 90th percentiles are considered as the lower and upper bounds, respectively, for each parameter.

**Table 1:** Rundown of the utilized datasets.

| Source | Year | Country | Number of fetuses | Weeks |
| --- | --- | --- | --- | --- |
| Araujo Júnior *et al.* <sup>[1]</sup> | 2014 | Brazil | 31476 | 18–38 |
| Brons *et al.* <sup>[2]</sup> | 1990 | Netherlands | 63 | 12–40 |
| Browne *et al.* <sup>[3]</sup> | 1992 | USA | 8285 | 10–44 |
| Buck Louis *et al.* <sup>[4]</sup> (Asian) | 2015 | USA | 460 | 10–40 |
| Buck Louis *et al.* <sup>[4]</sup> (Black) | 2015 | USA | 611 | 10–40 |
| Buck Louis *et al.* <sup>[4]</sup> (Hispanic) | 2015 | USA | 649 | 10–40 |
| Buck Louis *et al.* <sup>[4]</sup> (White) | 2015 | USA | 614 | 10–40 |
| Chitty *et al.* <sup>[8;7;9]</sup> | 1994 | UK | 594–649 | 12–42 |
| de la Vega *et al.* <sup>[11]</sup> | 2008 | Puerto Rico | 548 | 14–38 |
| Dubiel *et al.* <sup>[13]</sup> | 2008 | Poland | 959 | 20–42 |
| Dulger *et al.* <sup>[14]</sup> | 2024 | Turkey | 1132 | 15–40 |
| Fouad *et al.* <sup>[17]</sup> | 2021 | Egypt | 540 | 14–40 |
| Giorlandino *et al.* <sup>[19]</sup> | 2009 | Italy | 4896 | 14–41 |
| Johnsen *et al.* <sup>[25]</sup> | 2006 | Norway | 650 | 10–42 |
| Jung *et al.* <sup>[27]</sup> | 2007 | South Korea | 10455 | 12–40 |
| Kiserud *et al.* <sup>[28]</sup> | 2017 | Multinational | 1387 | 14–40 |
| Kwon *et al.* <sup>[29]</sup> | 2014 | South Korea | 986 | 15–40 |
| Lai and Yeo <sup>[30]</sup> | 1992 | Singapore | 6374 | 14–41 |
| Lessoway *et al.* <sup>[31]</sup> | 1998 | Canada | 1396 | 11–42 |
| Lindström *et al.* <sup>[32]</sup> | 2020 | Sweden | 583 | 12–42 |
| Munim *et al.* <sup>[35]</sup> | 2012 | Pakistan | 1228 | 20–39 |
| Paladini *et al.* <sup>[37]</sup> | 2005 | Italy | 626 | 16–40 |
| Papageorgiou *et al.* <sup>[38]</sup> | 2014 | Multinational | 13108 | 14–40 |
| Saksiriwuttho *et al.* <sup>[43]</sup> | 2007 | Thailand | 628 | 14–41 |
| Stampalija *et al.* <sup>[50]</sup> | 2020 | Italy | 7347 | 15–40 |
| Verburg *et al.* <sup>[54]</sup> | 2008 | Netherlands | 8313 | 12–40 |

### 2.4 Formulas

The formulas under investigation, which take into account more than one independent parameter, are presented in Table 2.

**Table 2:** List of the formulas under investigation.

| Source | Formula |
| --- | --- |
| Combs (1993)<sup>[10]</sup> | $0.23718 (AC)^2 (FL) + 0.03312 (HC)^3$ |
| Ferrero (1994)<sup>[16]</sup> | $10^{0.77125+0.13244(AC)-0.12996(FL)-1.73588(AC)^2/1000+3.09212(AC)(FL)/1000+2.18984(FL)/(AC)}$ |
| Hadlock I (1985)<sup>[20]</sup> | $10^{1.304+0.05281(AC)+0.1938(FL)-0.004(AC)(FL)}$ |
| Hadlock II (1985)<sup>[20]</sup> | $10^{1.335-0.0034(AC)(FL)+0.0316(BPD)+0.0457(AC)+0.1623(FL)}$ |
| Hadlock III (1985)<sup>[20]</sup> | $10^{1.326-0.00326(AC)(FL)+0.0107(HC)+0.0438(AC)+0.158(FL)}$ |
| Hadlock IV (1985)<sup>[20]</sup> | $10^{1.3596+0.00061(BPD)(AC)+0.424(AC)+0.174(FL)+0.0064(HC)-0.00386(AC)(FL)}$ |
| Halaska (2006)<sup>[21]</sup> | $10^{0.64041(BPD)-0.03257(BPD)^2+0.00154(AC)(FL)}$ |
| Hsieh (1987)<sup>[24]</sup> | $10^{2.1315+0.0056541(BPD)(AC)-0.00015515(BPD)(AC)^2+0.000019782(AC)^3+0.052594(BPD)}$ |
| INTERGROWTH-21 (2017)<sup>[51]</sup> | $e^{5.084820-54.06633((AC)/100)^3-95.80076((AC)/100)^3 \ln((AC)/100)+3.136370(HC)/100}$ |
| Jordaan I (1983)<sup>[26]</sup> | $10^{-1.1683+0.0377(AC)+0.095(BPD)-0.0015(BPD)(AC)}$ |
| Jordaan II (1983)<sup>[26]</sup> | $10^{0.9119+0.0488(HC)+0.0824(AC)+0.001599(AC)(HC)}$ |
| Merz (1988)<sup>[33]</sup> | $3200.40479 + 157.07186 (AC) + 15.90391 (BPD)^2$ |
| Ott (1986)<sup>[36]</sup> | $10^{-2.0661+0.04355(HC)+0.05394(AC)-0.0008582(AC)(HC)+1.2594(AC)(FL)(AC)}$ |
| Roberts (1985)<sup>[42]</sup> | $10^{1.6758+0.01707(AC)+0.042478(BPD)+0.05216(FL)+0.01604(HC)}$ |
| Schild (2004)<sup>[45]</sup> | $5381.193 + 150.324 (HC) + 2.069 (FL)^3 + 0.0232 (AC)^3 - 6235.478 \log(HC)$ |
| Shepard I (1982)<sup>[47]</sup> | $10^{-1.599+0.144(BPD)+0.032(AC)-0.000111(AC)(BPD)^2}$ |
| Shepard II (1982)<sup>[47]</sup> | $10^{-1.7492+0.166(BPD)+0.046(AC)-0.002646(BPD)(AC)}$ |
| Thurnau (1983)<sup>[52]</sup> | $9.337 (BPD) (AC) - 229$ |
| Vintzileos I (1987)<sup>[55]</sup> | $10^{1.879+0.084(BPD)+0.026(AC)}$ |
| Vintzileos II (1987)<sup>[55]</sup> | $10^{2.082+0.004(HC)(FL)+0.0018(AC)-0.00000001509((HC)(FL))^3}$ |
| Warsof (1977)<sup>[56]</sup> | $10^{-1.599+0.144(BPD)+0.032(AC)-0.000111(BPD)^2(AC)}$ |
| Weiner I (1985)<sup>[57]</sup> | $10^{1.6961+0.02253(HC)+0.01645(AC)+0.06439(FL)}$ |
| Weiner II (1985)<sup>[57]</sup> | $10^{1.6575+0.04035(HC)+0.01285(AC)}$ |
| Woo (1986)<sup>[58]</sup> | $1.4 (BPD) (AC) (HC) - 200$ |
| Woo I (1985)<sup>[59]</sup> | $10^{1.54+0.15(BPD)+0.00111(AC)^2-0.0000764(BPD)(AC)^2+0.05(FL)-0.000992(AC)(FL)}$ |
| Woo II (1985)<sup>[59]</sup> | $10^{1.14+0.16(BPD)+0.05(AC)-2.8(BPD)(AC)/1000+0.04(FL)-4.9(AC)(FL)/10000}$ |
| Woo III (1985)<sup>[59]</sup> | $10^{1.13+0.18(BPD)+0.05(AC)-3.35(BPD)(AC)/1000}$ |
| Woo IV (1985)<sup>[59]</sup> | $10^{1.63+0.16(BPD)+0.00111(AC)^2-0.0000859(BPD)(AC)^2}$ |
| Woo V (1985)<sup>[59]</sup> | $10^{0.59+0.08(AC)+0.28(FL)-0.00716(AC)(FL)}$ |

### 2.5 Estimation of the mean of indices with bootstrapping

The mean values of the indices of each formula are estimated using non-parametric bootstrapping, which is described in Casella and Berger^[6]^. This method is chosen due to its no distributional assumptions, robustness to outliers, and usefulness in small samples.

To obtain the median, along with a 95% confidence interval (CI) for the estimation of the mean of the Sobol’ indices, the methodology outlined below is employed.

1. The Sobol’ indices are calculated for every combination of formula and dataset for each week in which data are available.
2. The calculated Sobol’ indices are grouped together by formula and gestation age (excluding weeks 43 and 44 due to scarcity of available data).
3. 10^6^ bootstrap samples are created for each aforementioned group
4. The average is calculated for each bootstrap sample, resulting in samples of size 10^6^ for the respective mean value of each group.
5. The median, along with a 95% CI, is estimated for each mean-value sample, using the empirical distribution function.

## 3 Results

Figures 3 to 31 present an estimate of the first-order Sobol’s indices for each parameter of every formula of Table 2, based on the datasets of Table 1. Each figure consists of an error bar for each parameter and for each gestation week. The median of the mean value of the indices is represented by a dot, while the whiskers represent a 95% CI. In addition, each figure displays the indices for each individual dataset as a scatter plot with a low alpha value.

**Figure 3:**
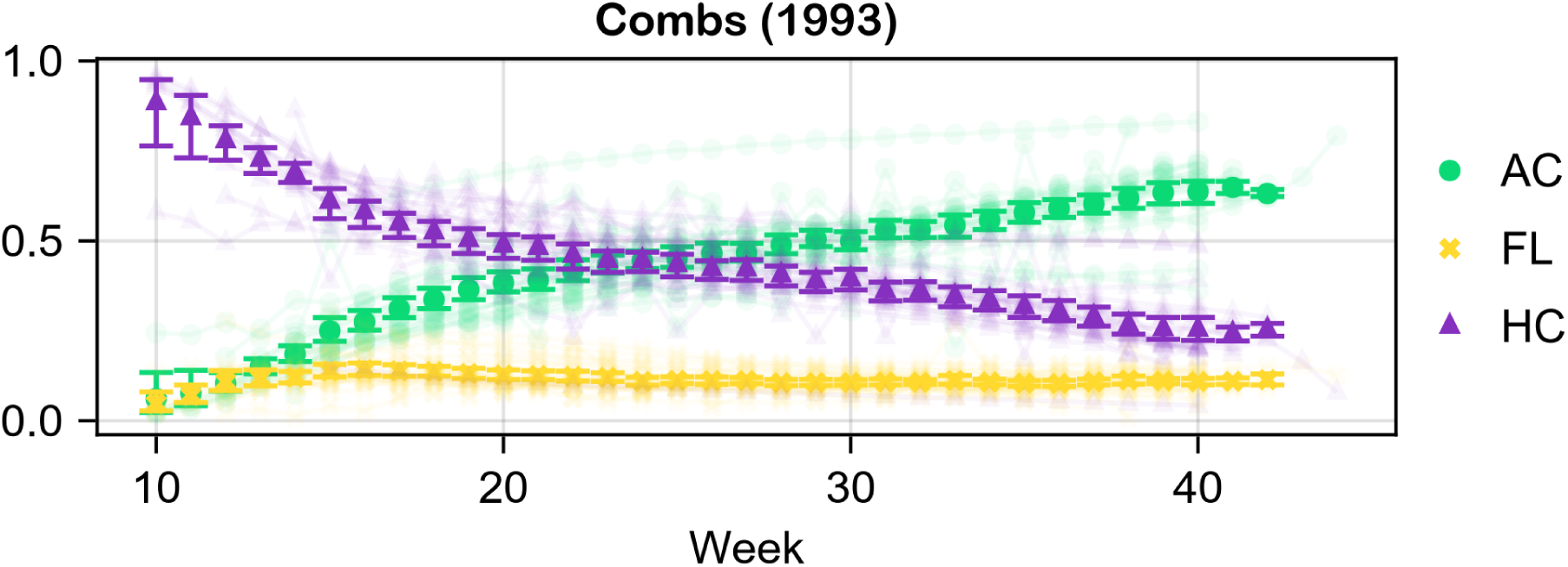
Estimate of the mean of the first-order Sobol’ indices of Combs (1993).

**Figure 4:**
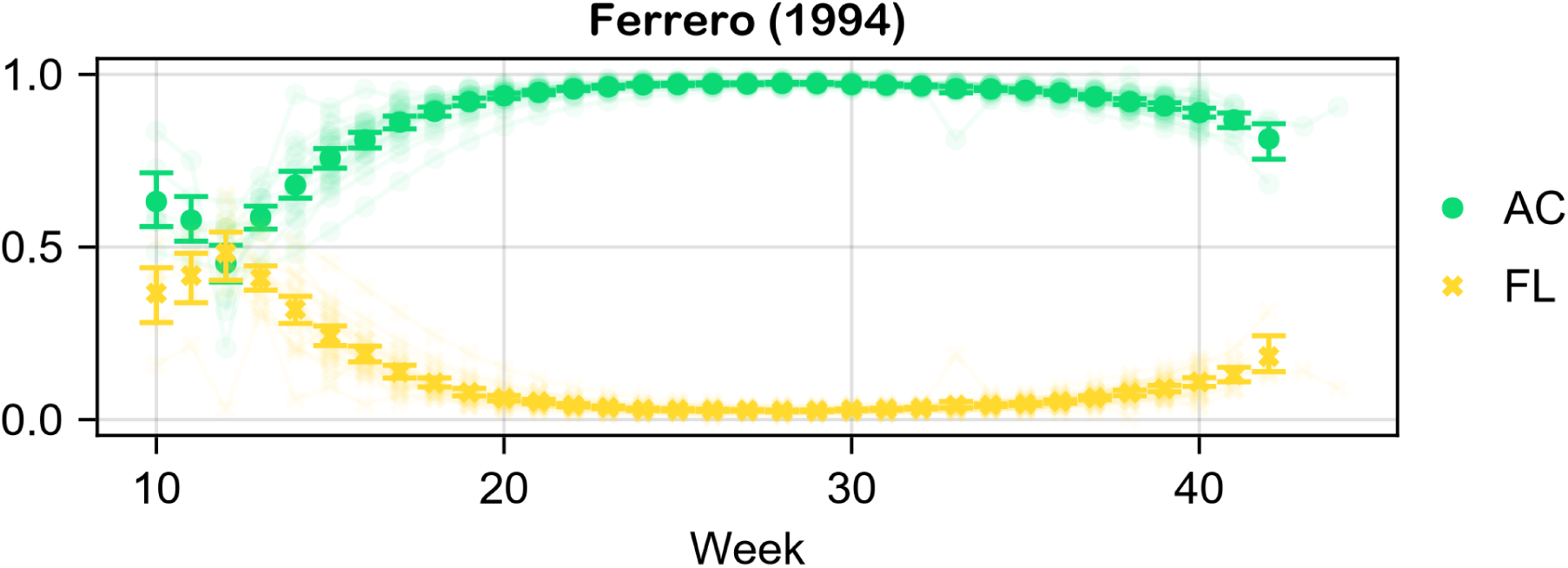
Estimate of the mean of the first-order Sobol’ indices of Ferrero (1994).

**Figure 5:**
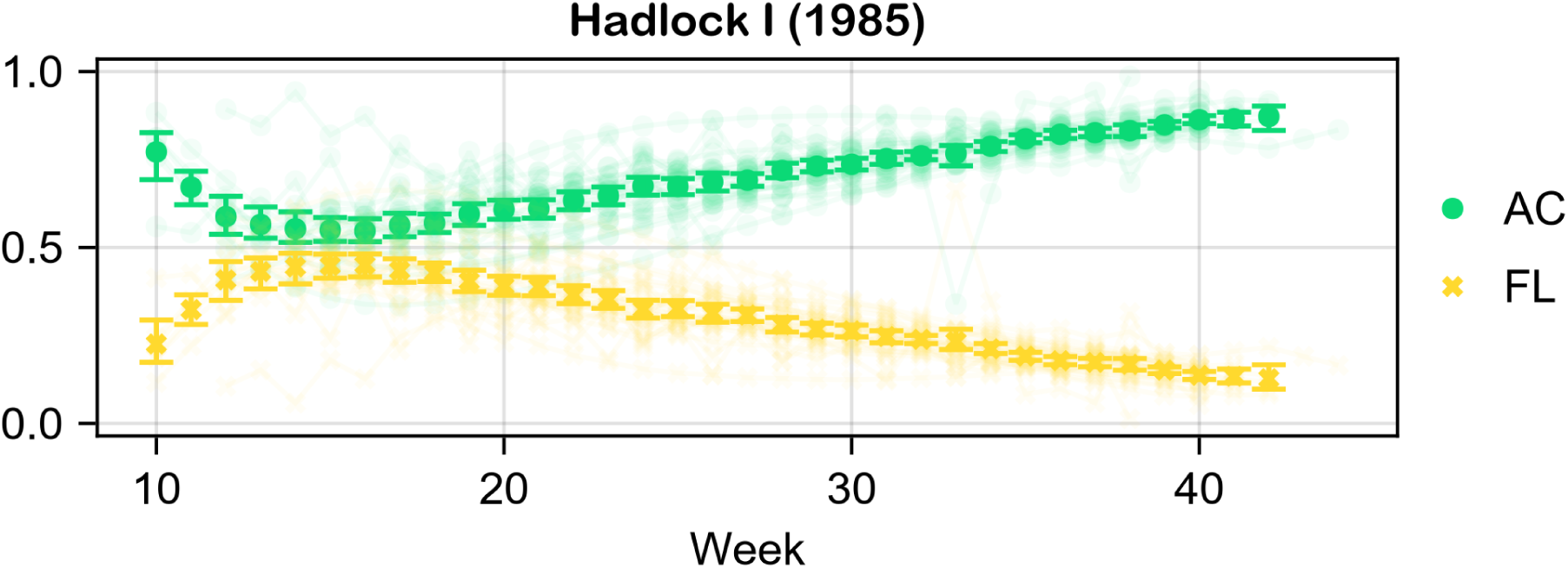
Estimate of the mean of the first-order Sobol’ indices of Hadlock I (1985).

**Figure 6:**
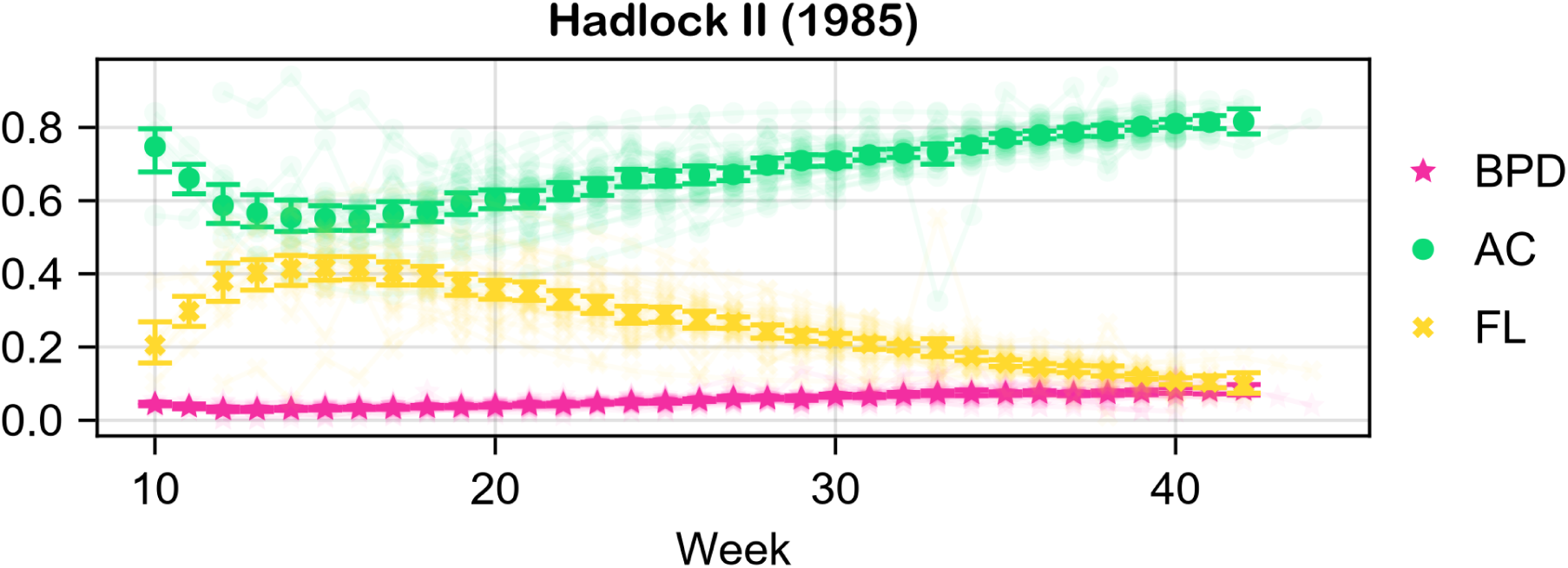
Estimate of the mean of the first-order Sobol’ indices of Hadlock II (1985).

**Figure 7:**
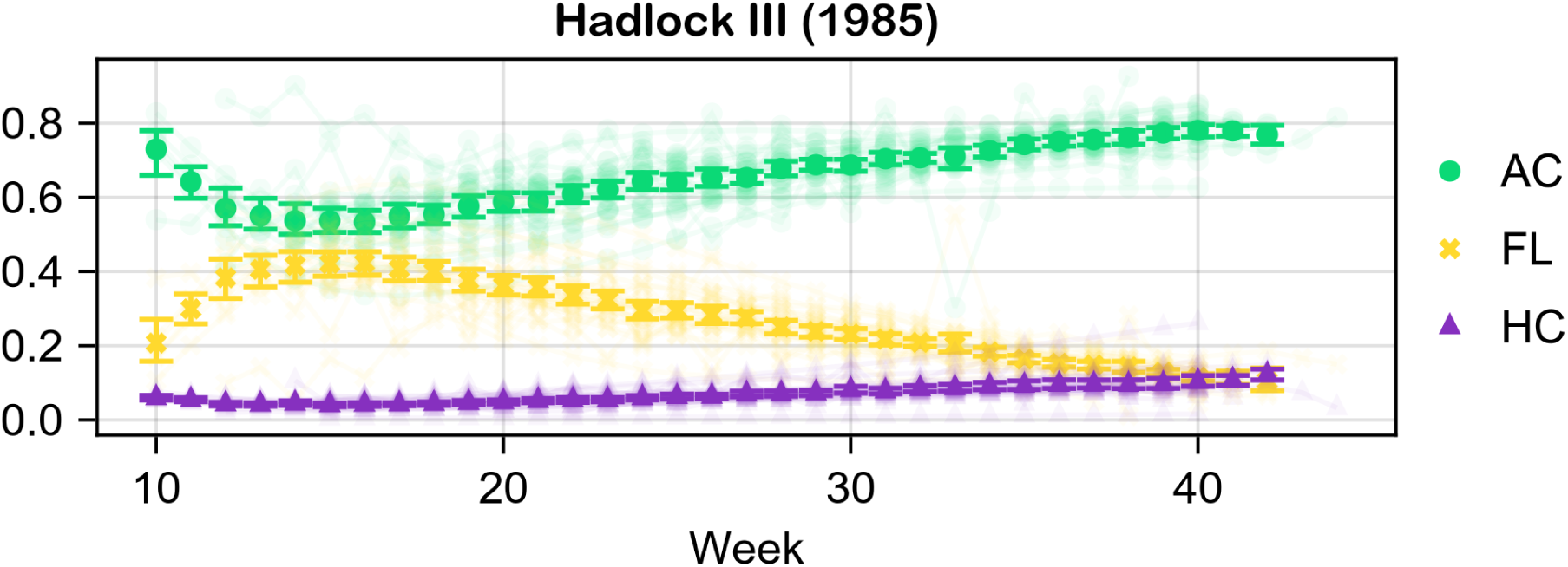
Estimate of the mean of the first-order Sobol’ indices of Hadlock III (1985).

**Figure 8:**
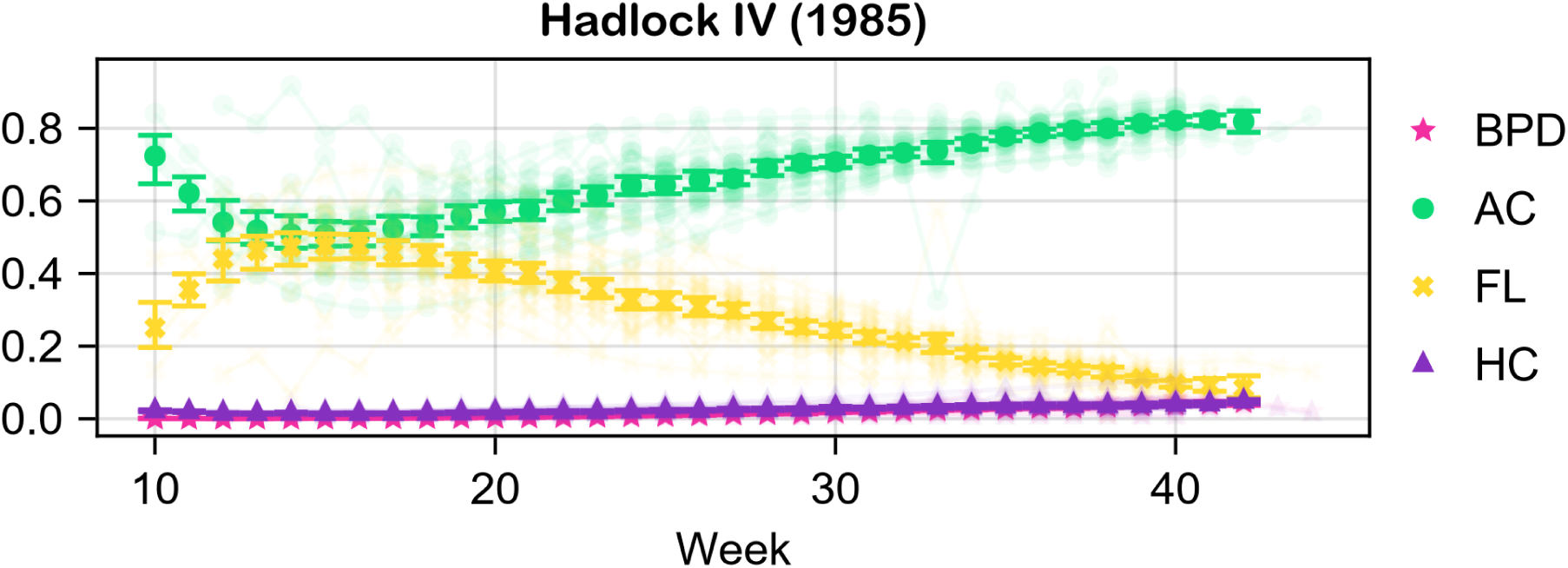
Estimate of the mean of the first-order Sobol’ indices of Hadlock IV (1985).

**Figure 9:**
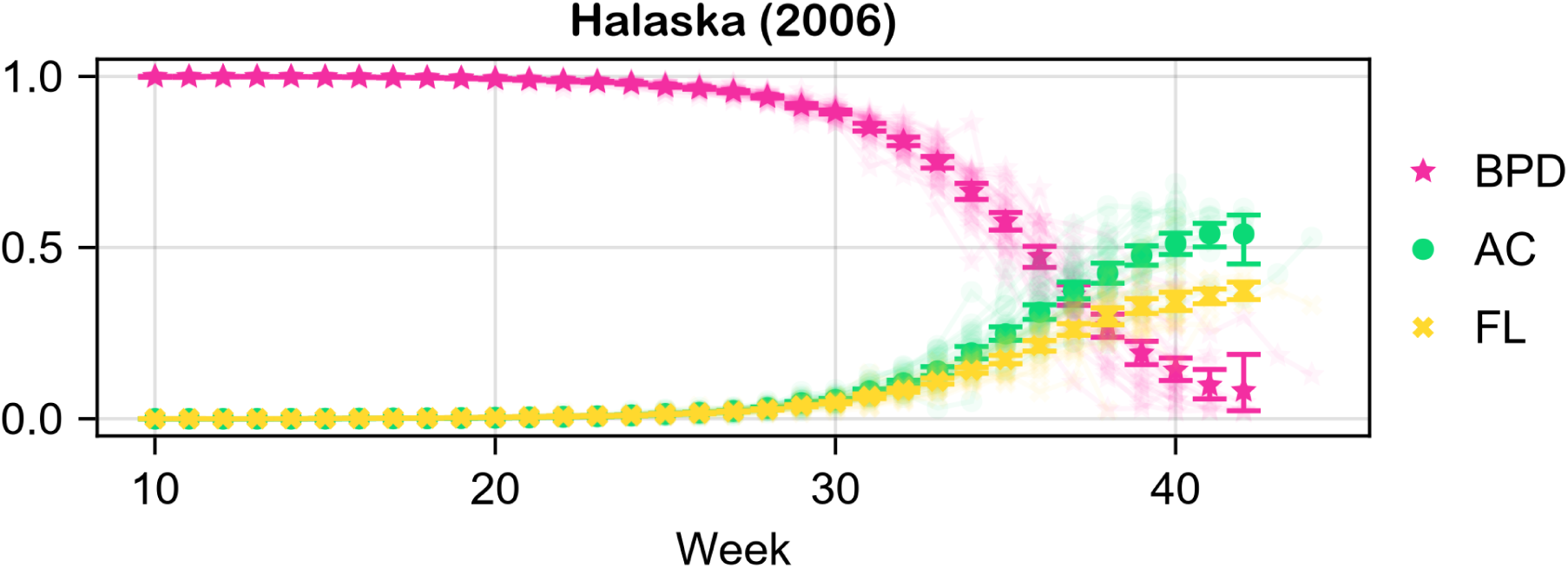
Estimate of the mean of the first-order Sobol’ indices of Halaska (2006).

**Figure 10:**
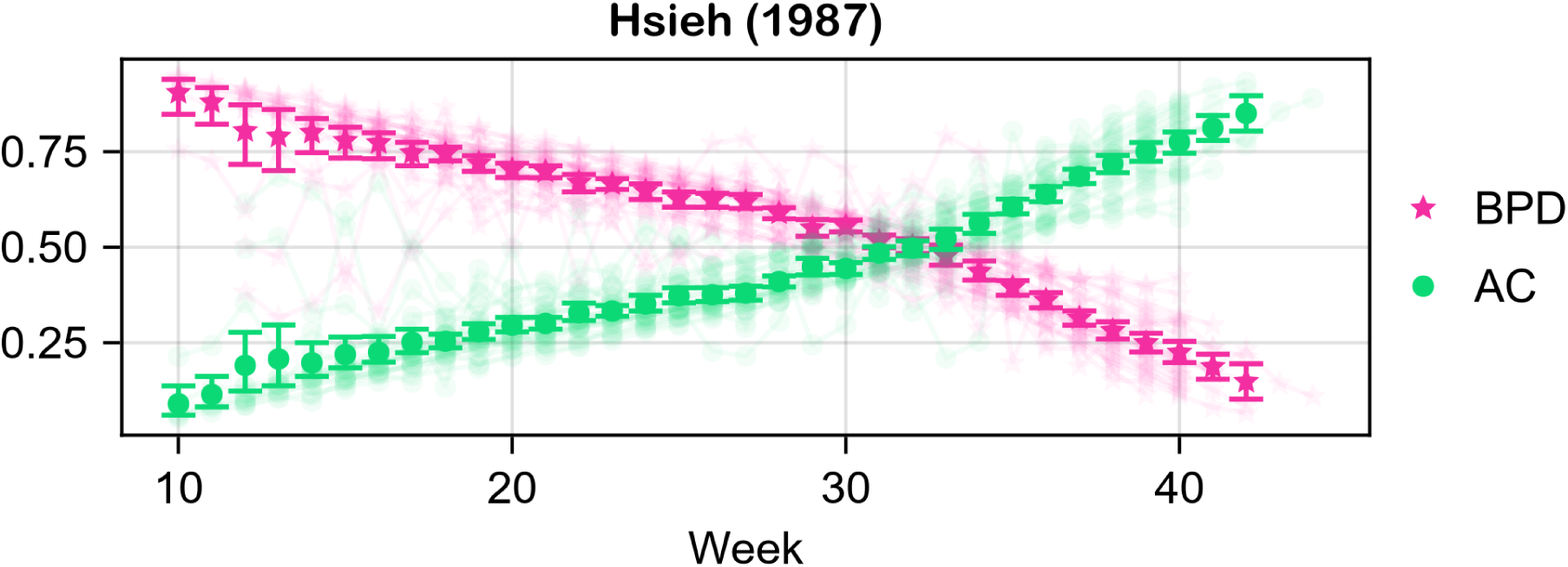
Estimate of the mean of the first-order Sobol’ indices of Hsieh (1987).

**Figure 11:**
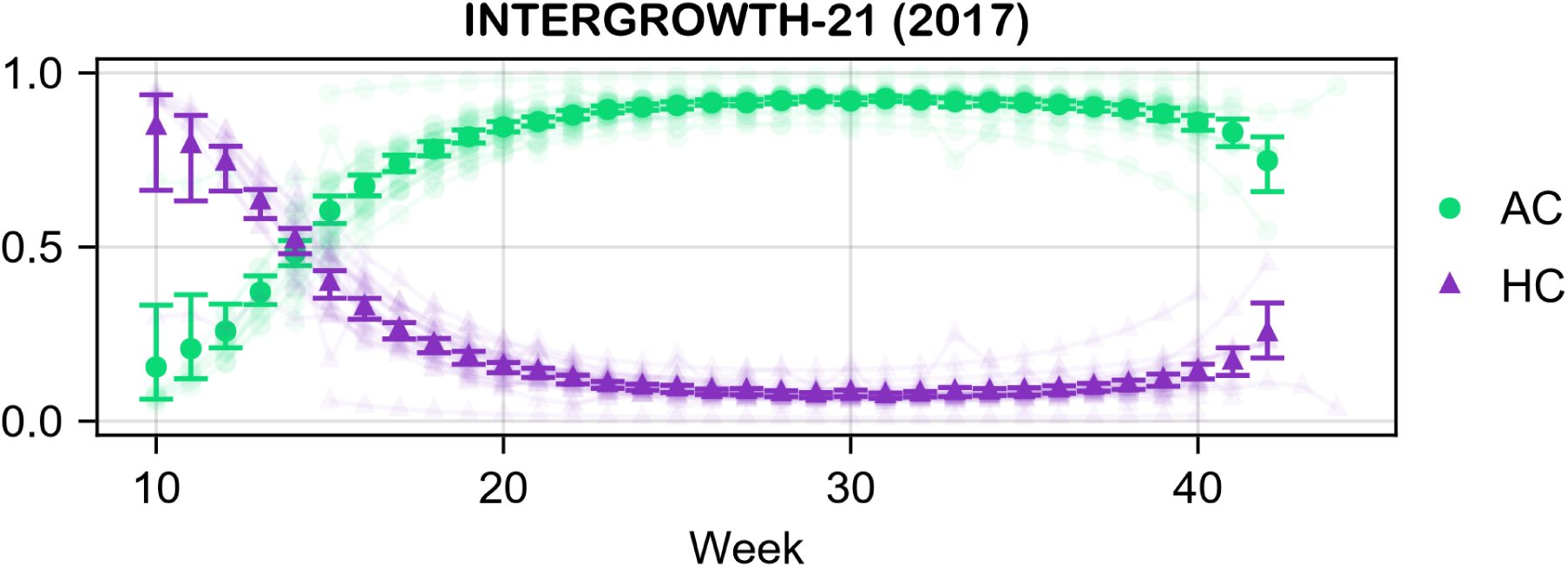
Estimate of the mean of the first-order Sobol’ indices of INTERGROWTH-21 (2017).

**Figure 12:**
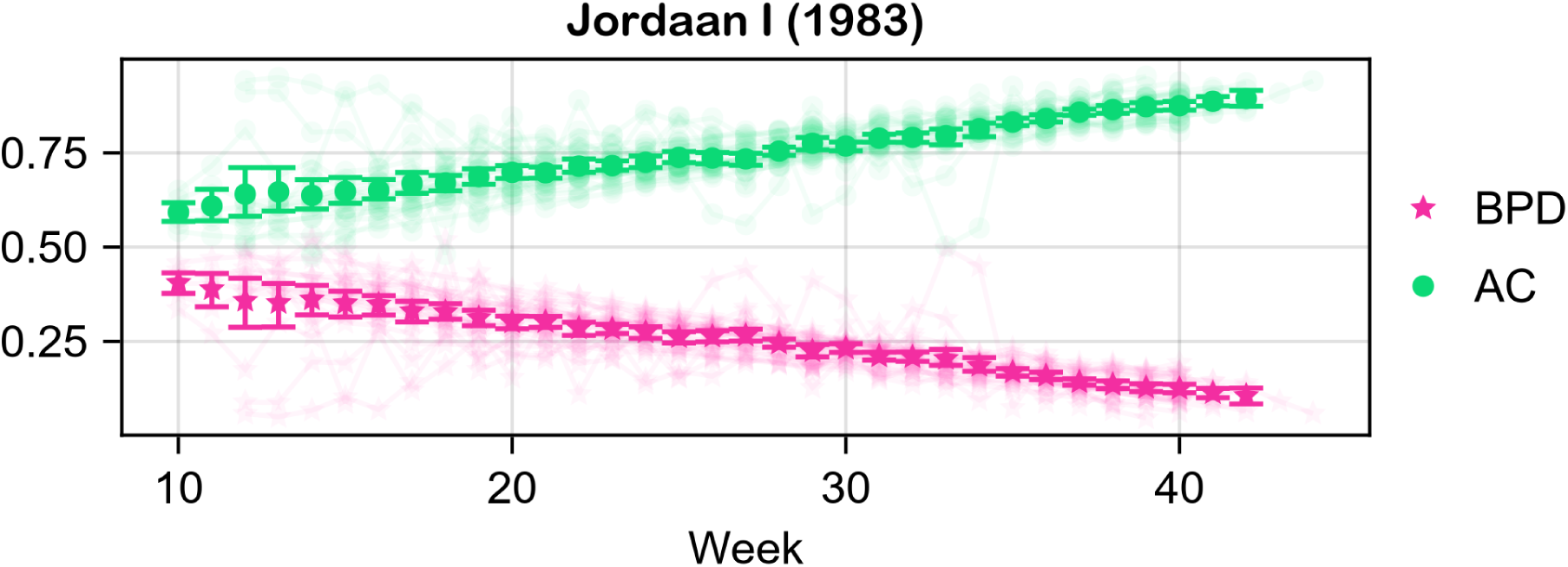
Estimate of the mean of the first-order Sobol’ indices of Jordaan I (1983).

**Figure 13:**
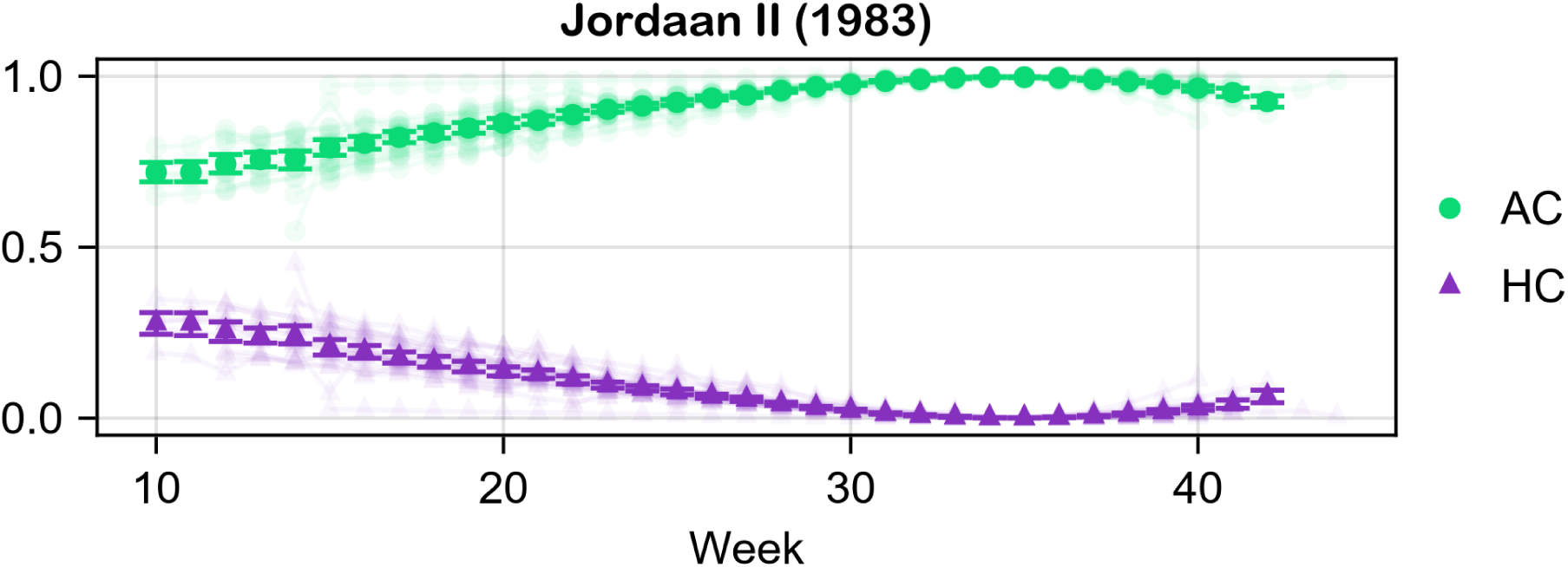
Estimate of the mean of the first-order Sobol’ indices of Jordaan II (1983).

**Figure 14:**
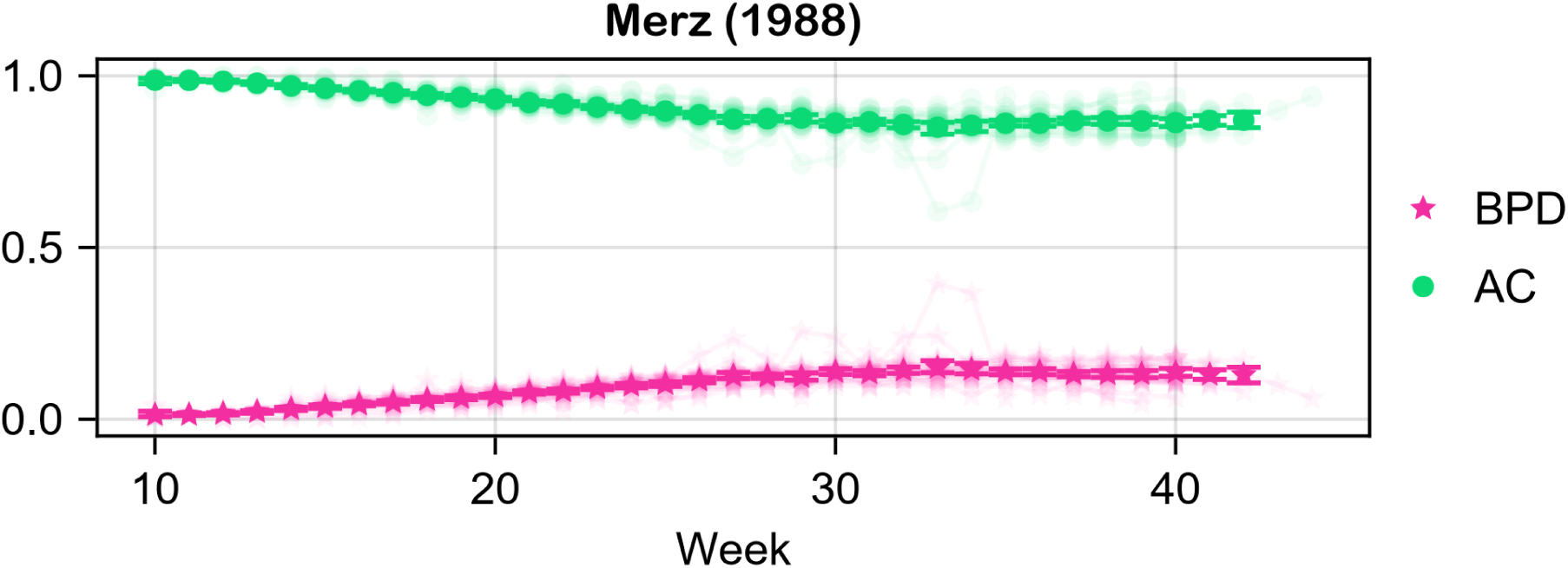
Estimate of the mean of the first-order Sobol’ indices of Merz (1988).

**Figure 15:**
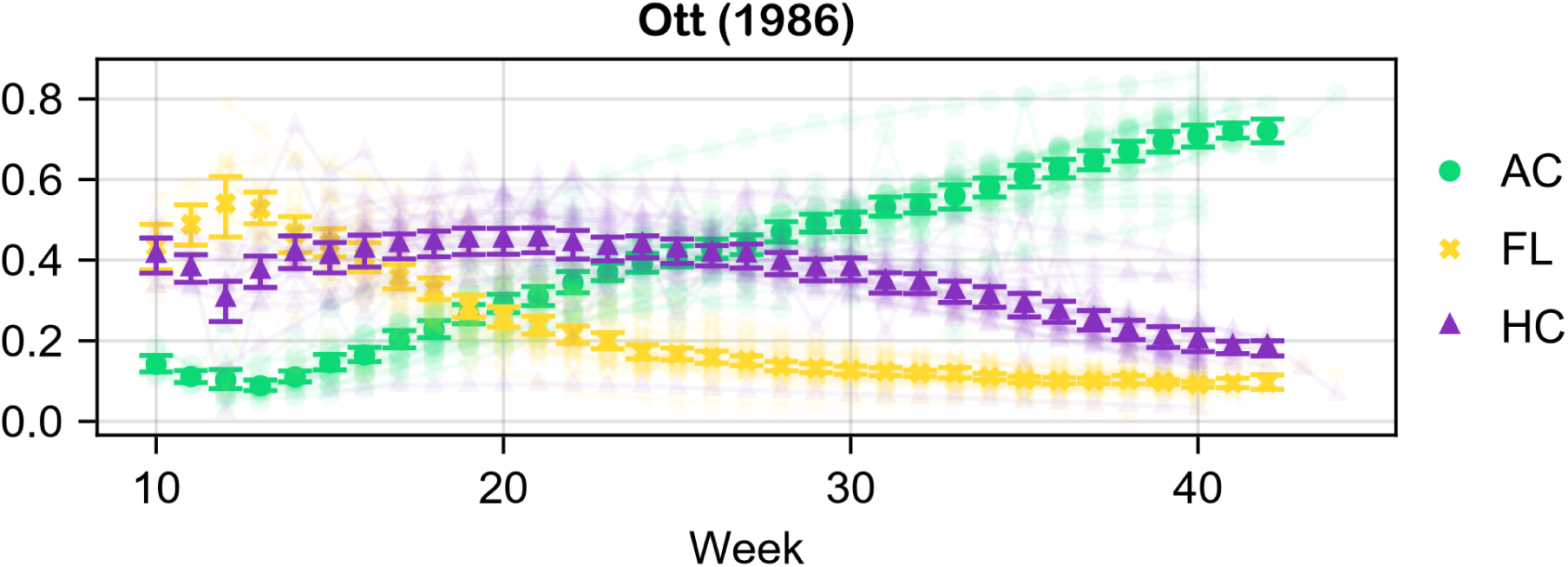
Estimate of the mean of the first-order Sobol’ indices of Ott (1986).

**Figure 16:**
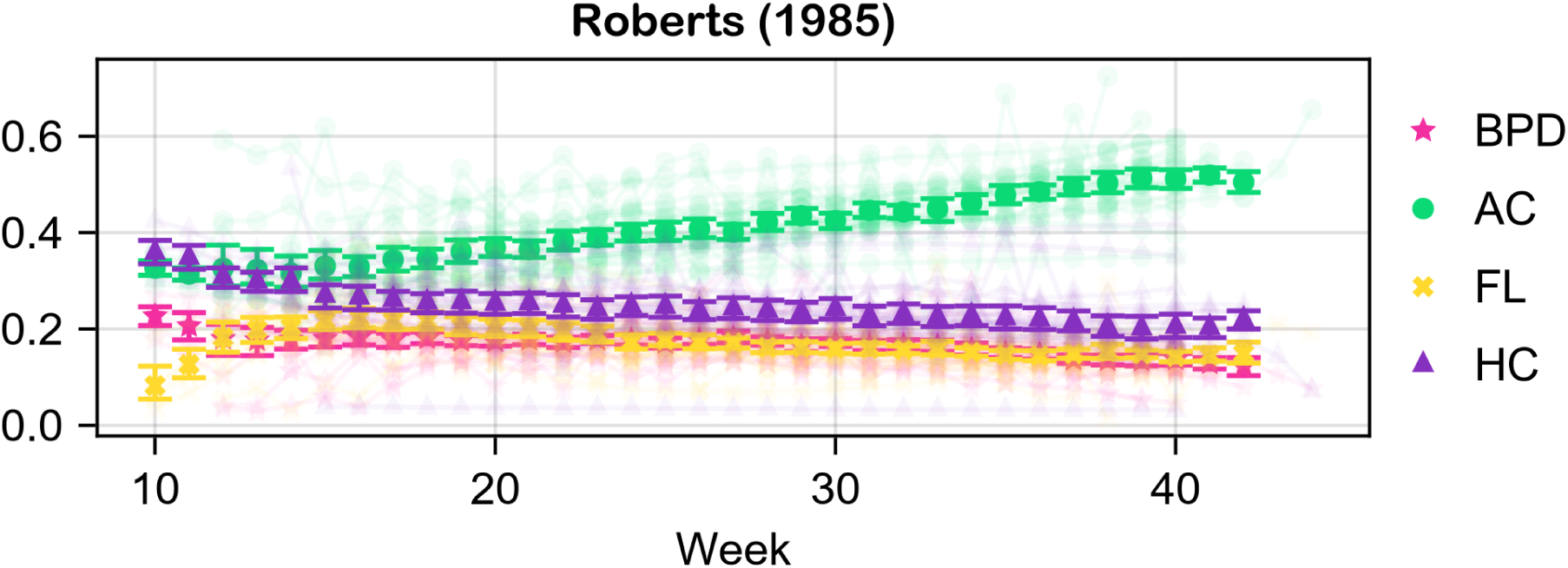
Estimate of the mean of the first-order Sobol’ indices of Roberts (1985).

**Figure 17:**
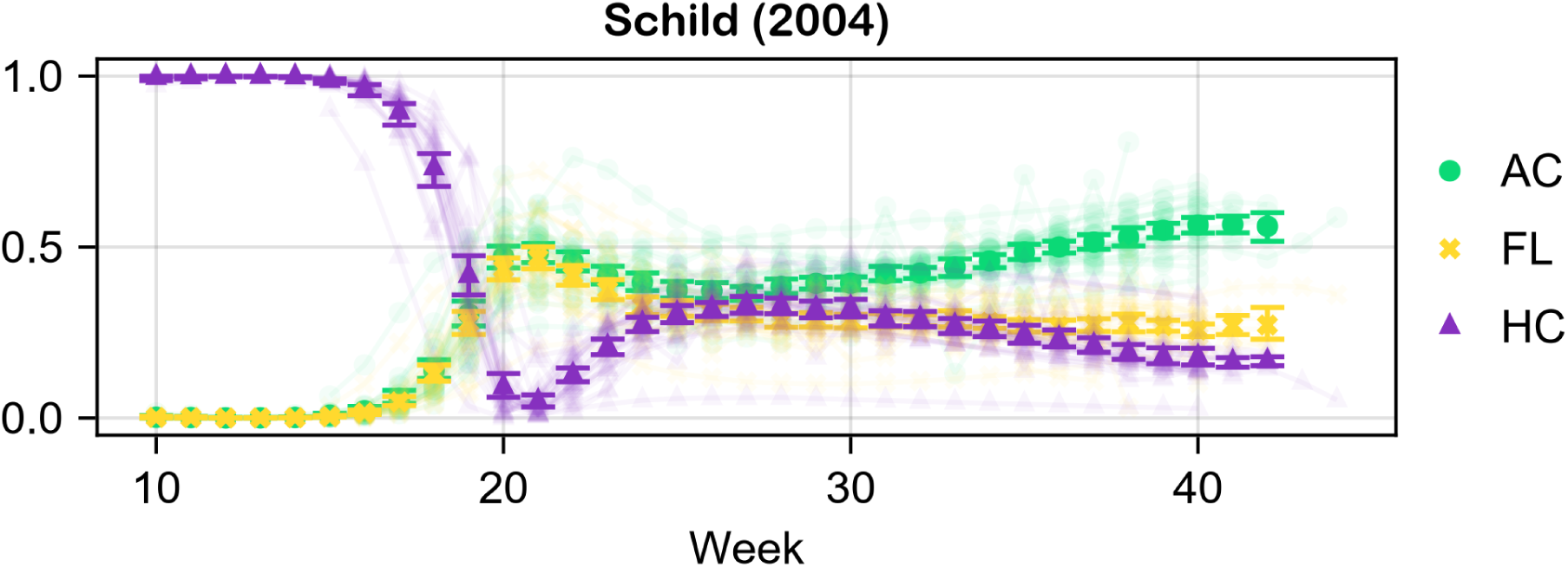
Estimate of the mean of the first-order Sobol’ indices of Schild (2004).

**Figure 18:**
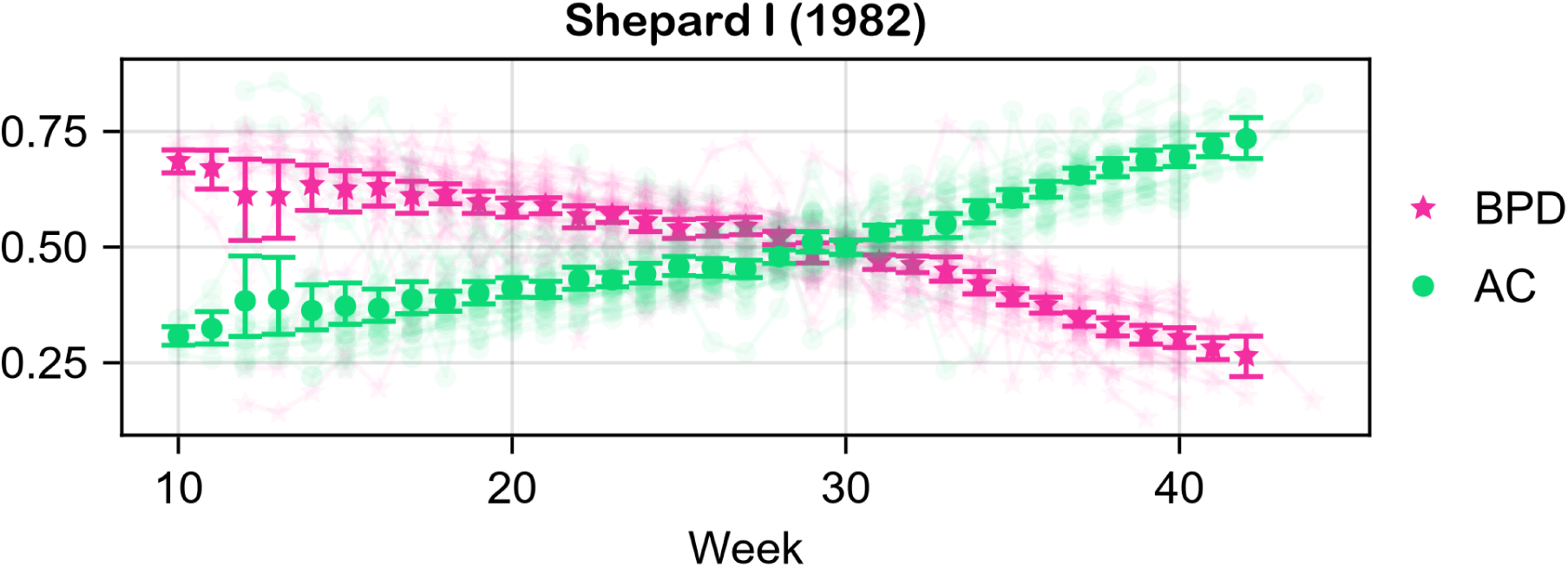
Estimate of the mean of the first-order Sobol’ indices of Shepard I (1982).

**Figure 19:**
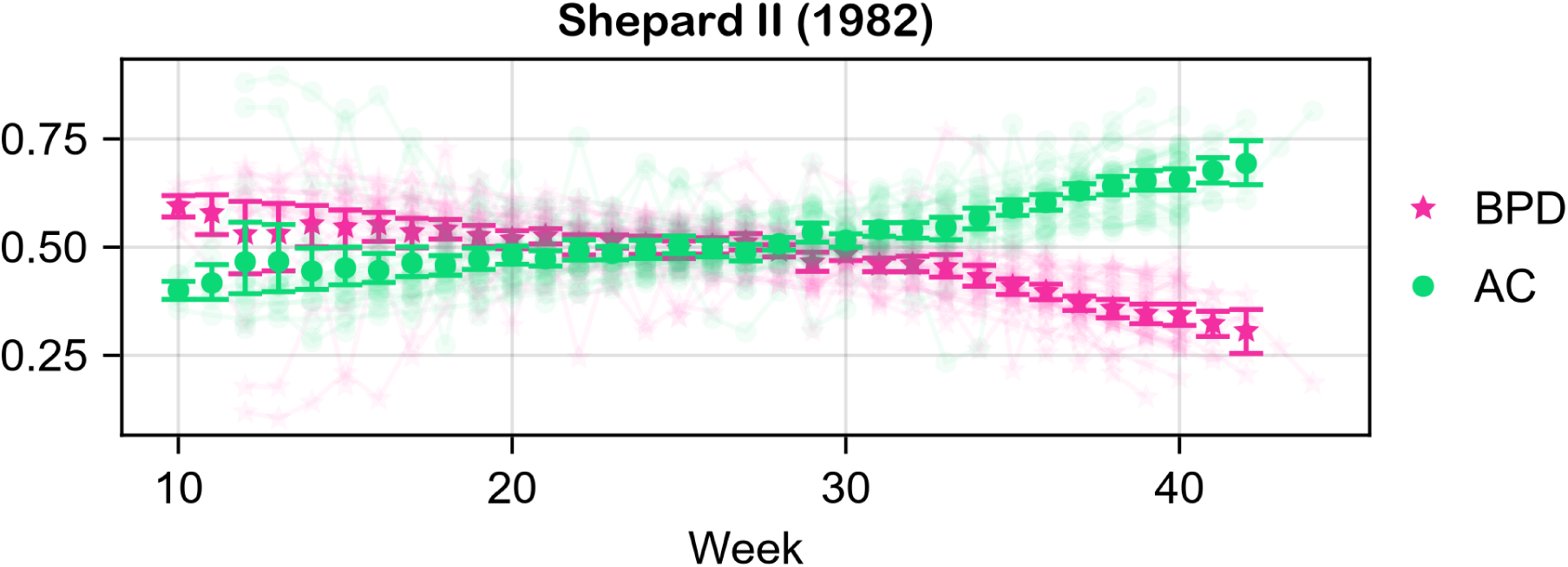
Estimate of the mean of the first-order Sobol’ indices of Shepard II (1982).

**Figure 20:**
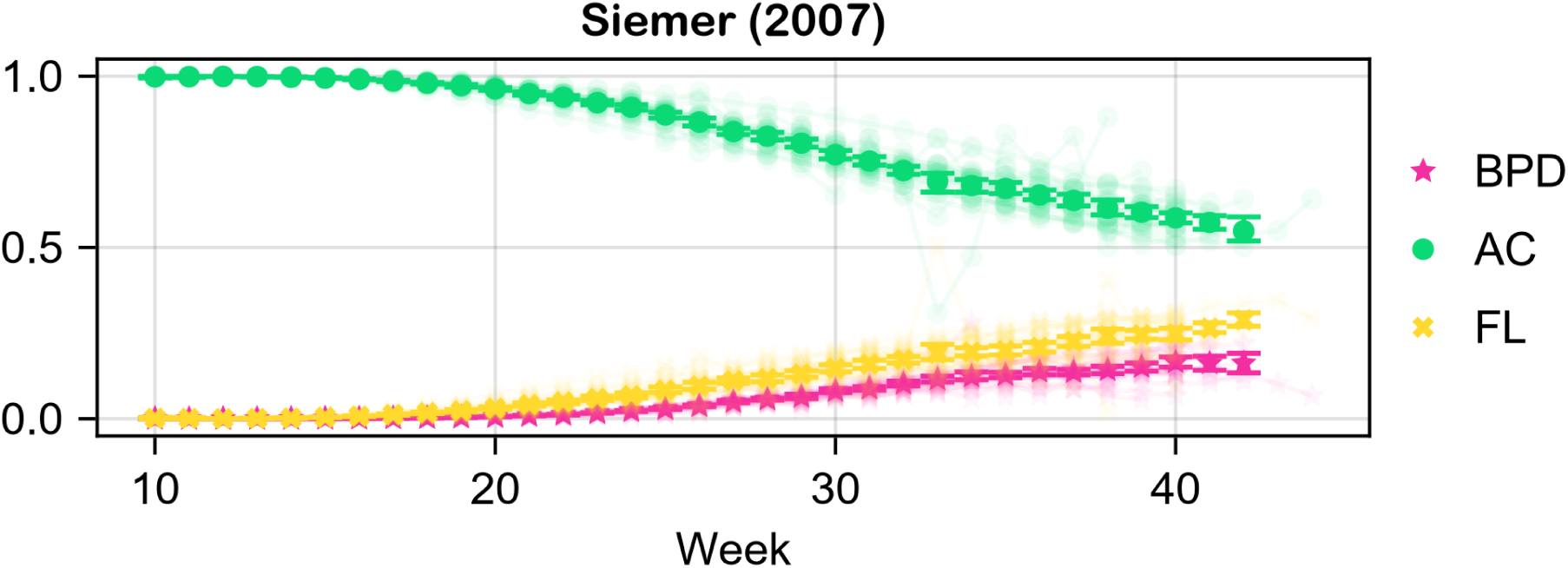
Estimate of the mean of the first-order Sobol’ indices of Siemer (2007).

**Figure 21:**
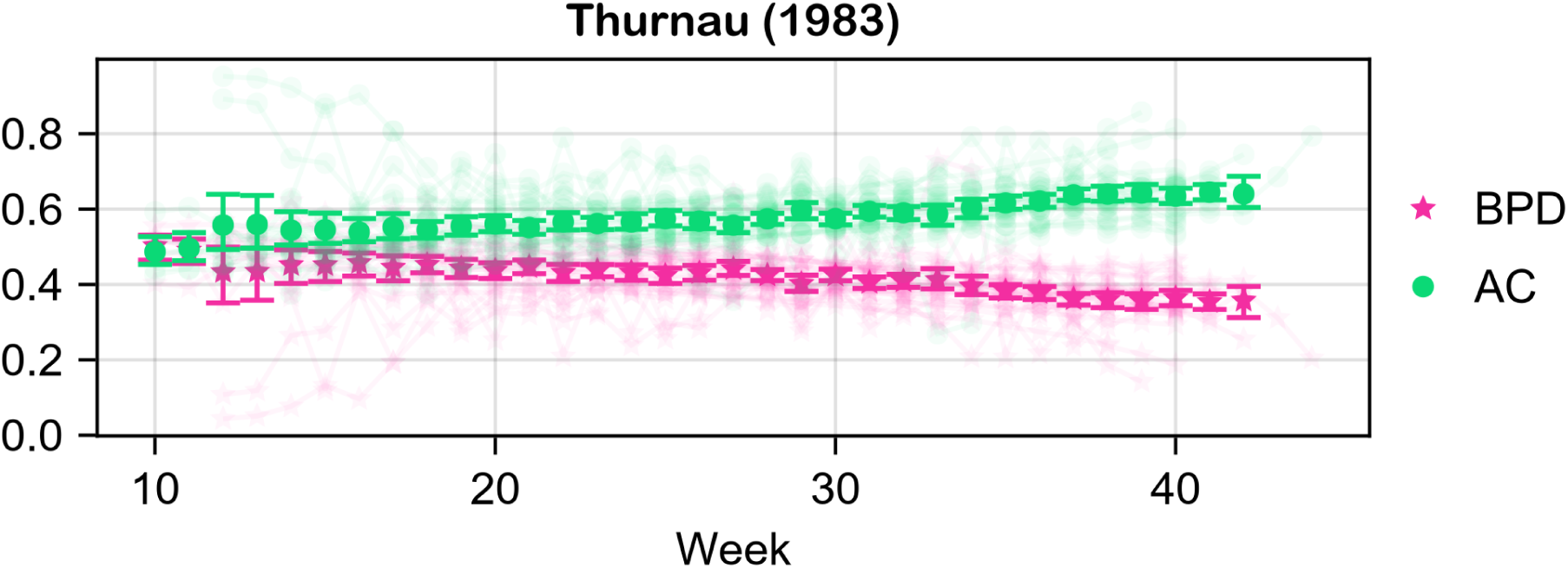
Estimate of the mean of the first-order Sobol’ indices of Thurnau (1983).

**Figure 22:**
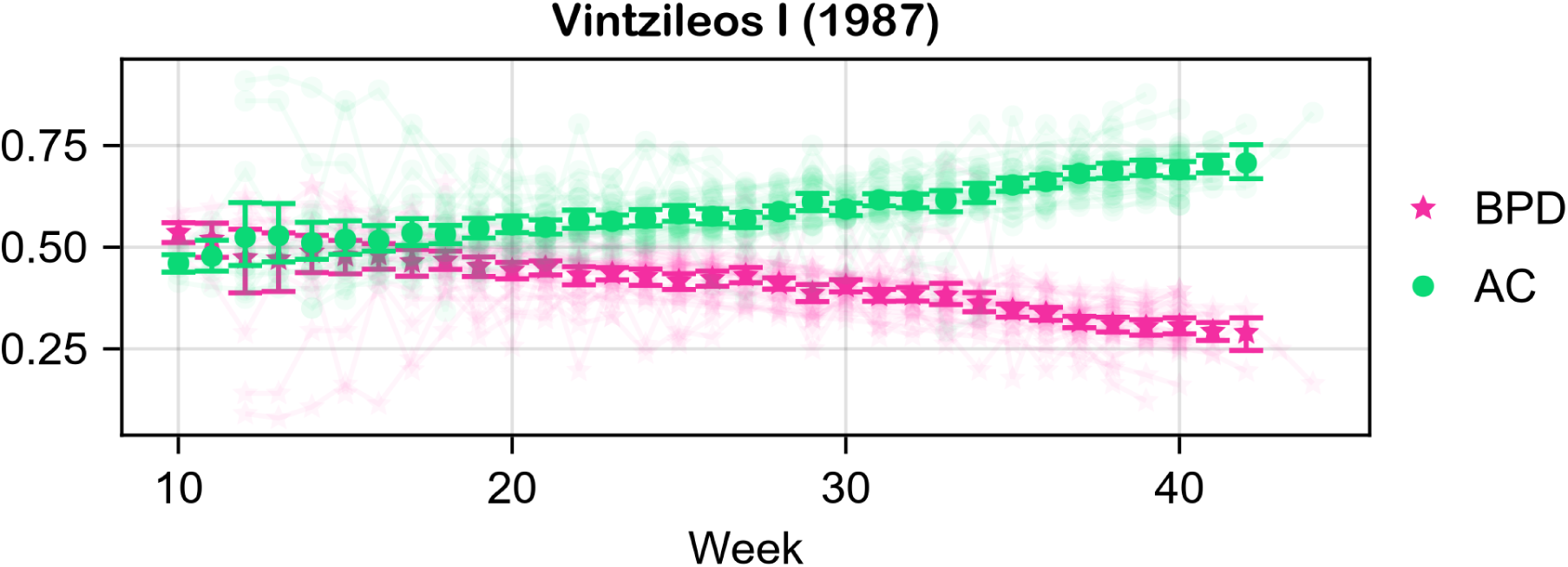
Estimate of the mean of the first-order Sobol’ indices of Vintzileos I (1987).

**Figure 23:**
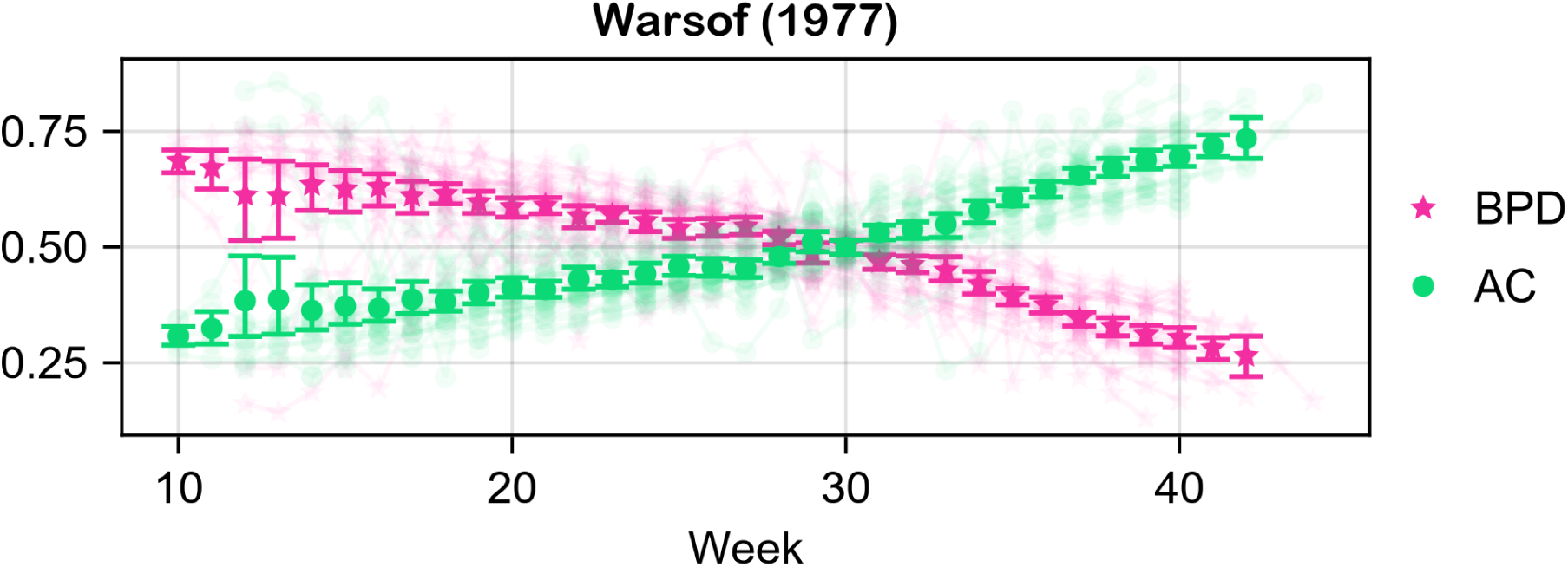
Estimate of the mean of the first-order Sobol’ indices of Warsof (1977).

**Figure 24:**
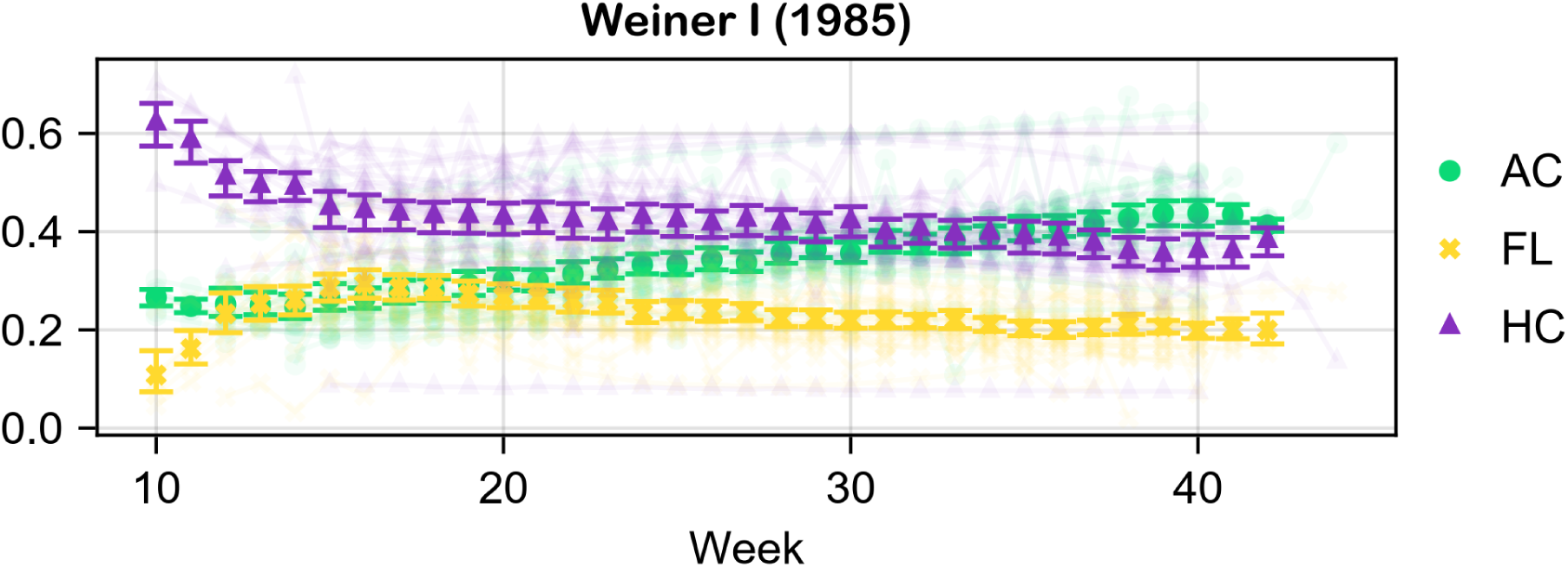
Estimate of the mean of the first-order Sobol’ indices of Weiner I (1985).

**Figure 25:**
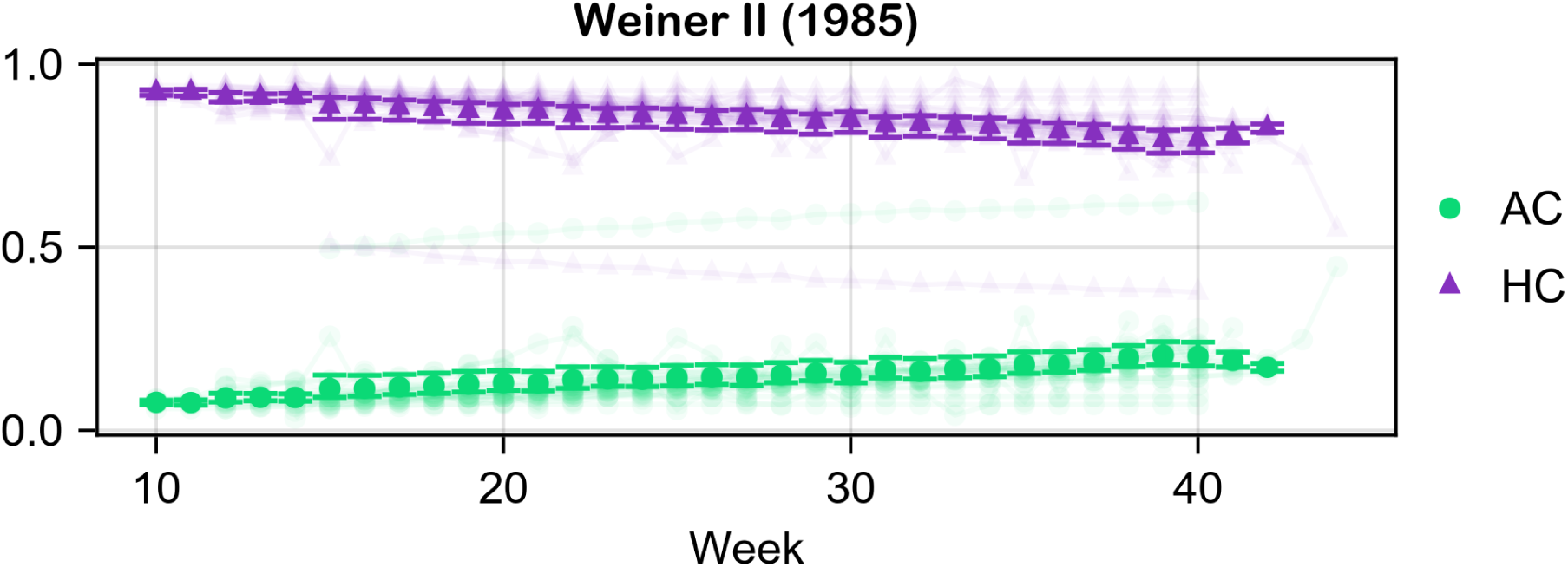
Estimate of the mean of the first-order Sobol’ indices of Weiner II (1985).

**Figure 26:**
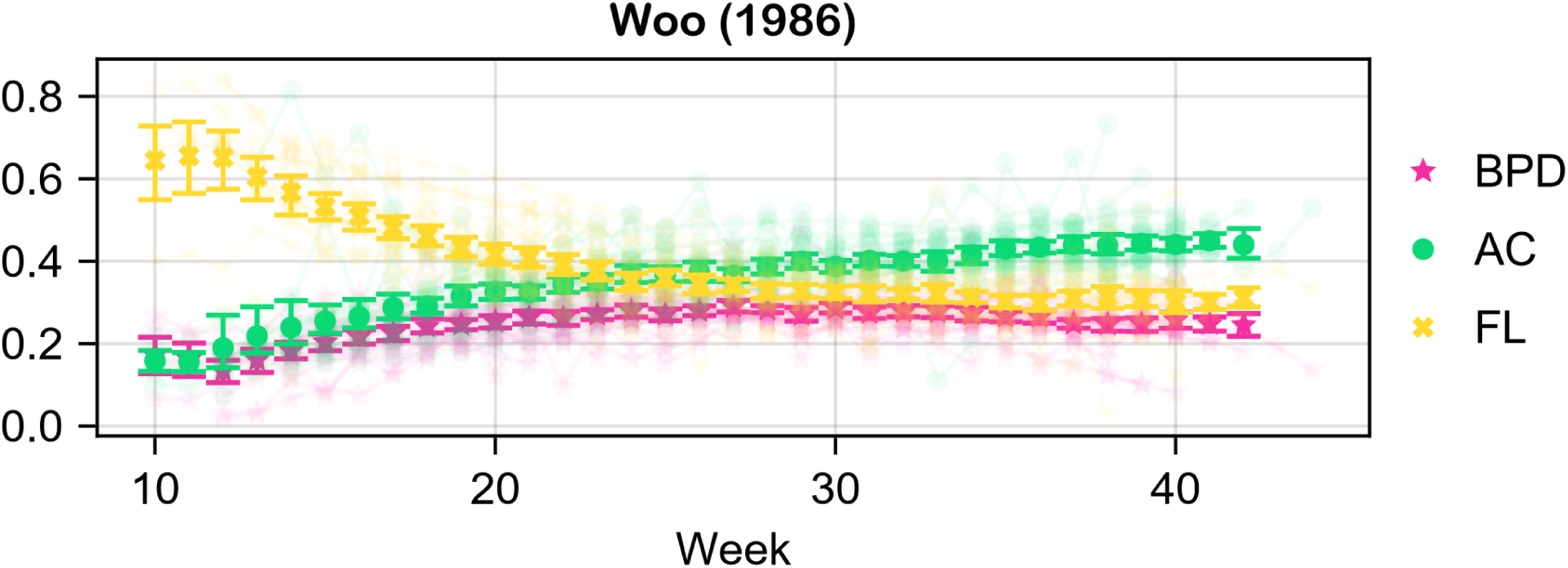
Estimate of the mean of the first-order Sobol’ indices of Woo (1986).

**Figure 27:**
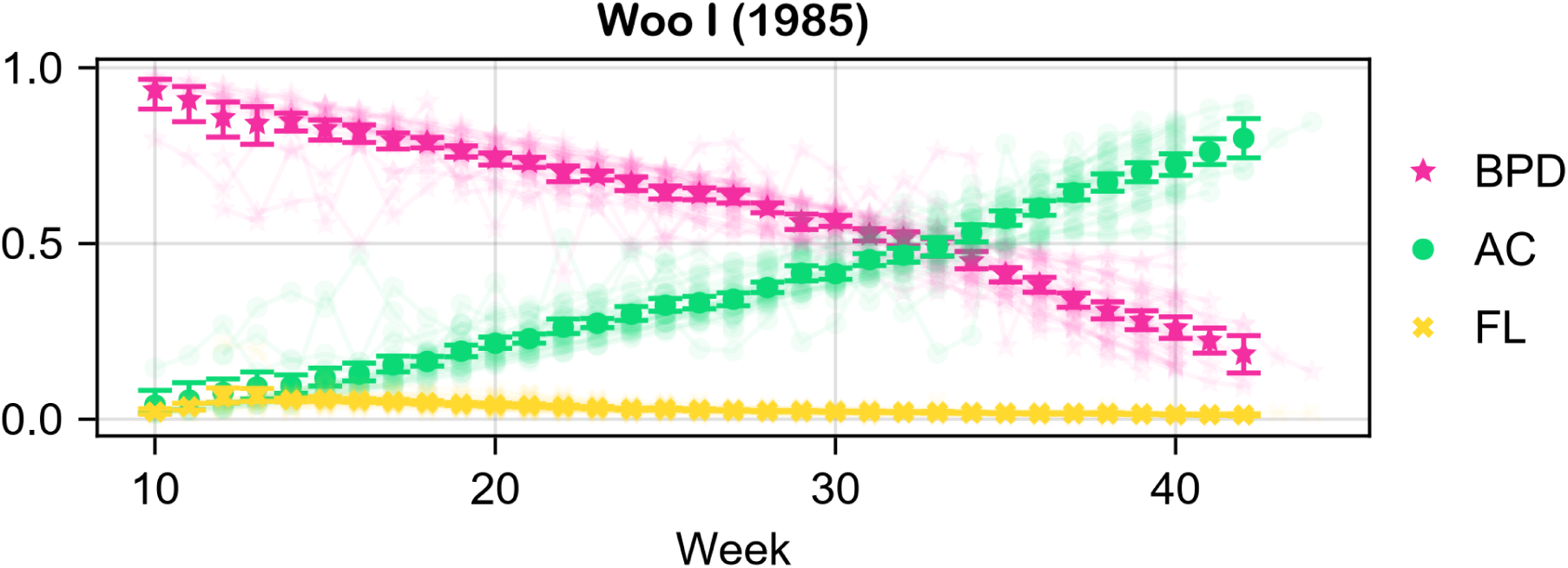
Estimate of the mean of the first-order Sobol’ indices of Woo I (1985).

**Figure 28:**
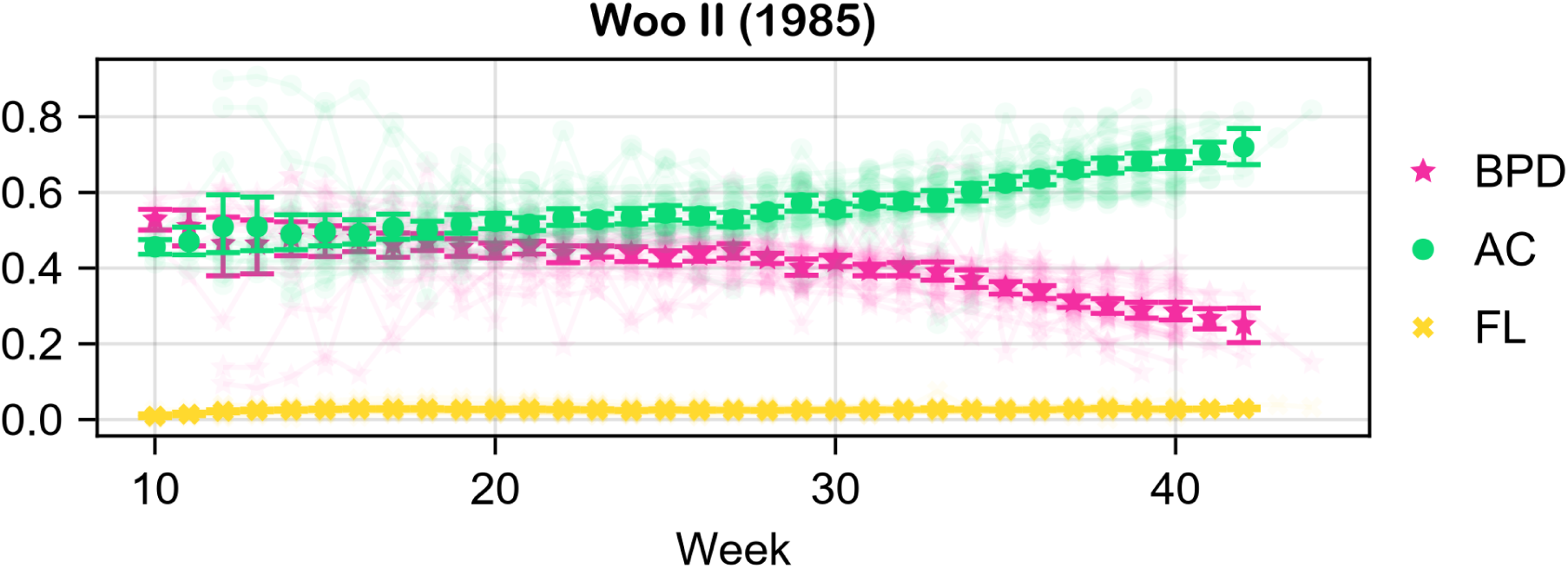
Estimate of the mean of the first-order Sobol’ indices of Woo II (1985).

**Figure 29:**
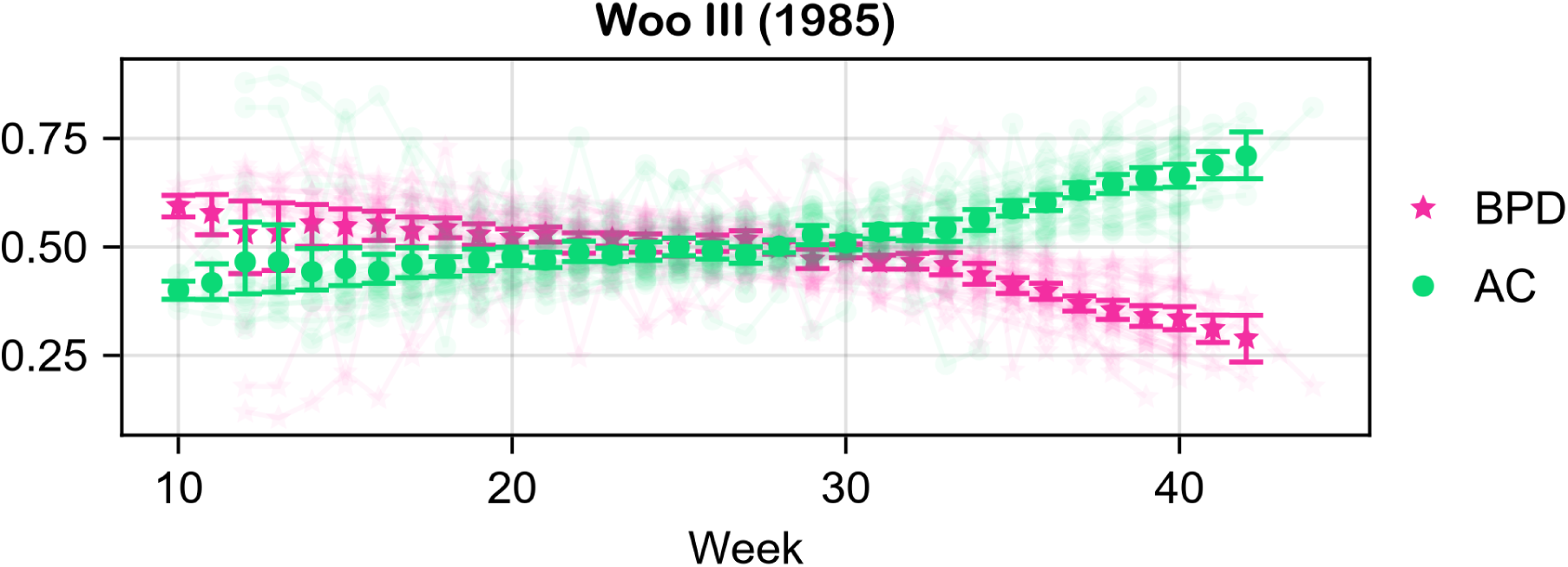
Estimate of the mean of the first-order Sobol’ indices of Woo III (1985).

**Figure 30:**
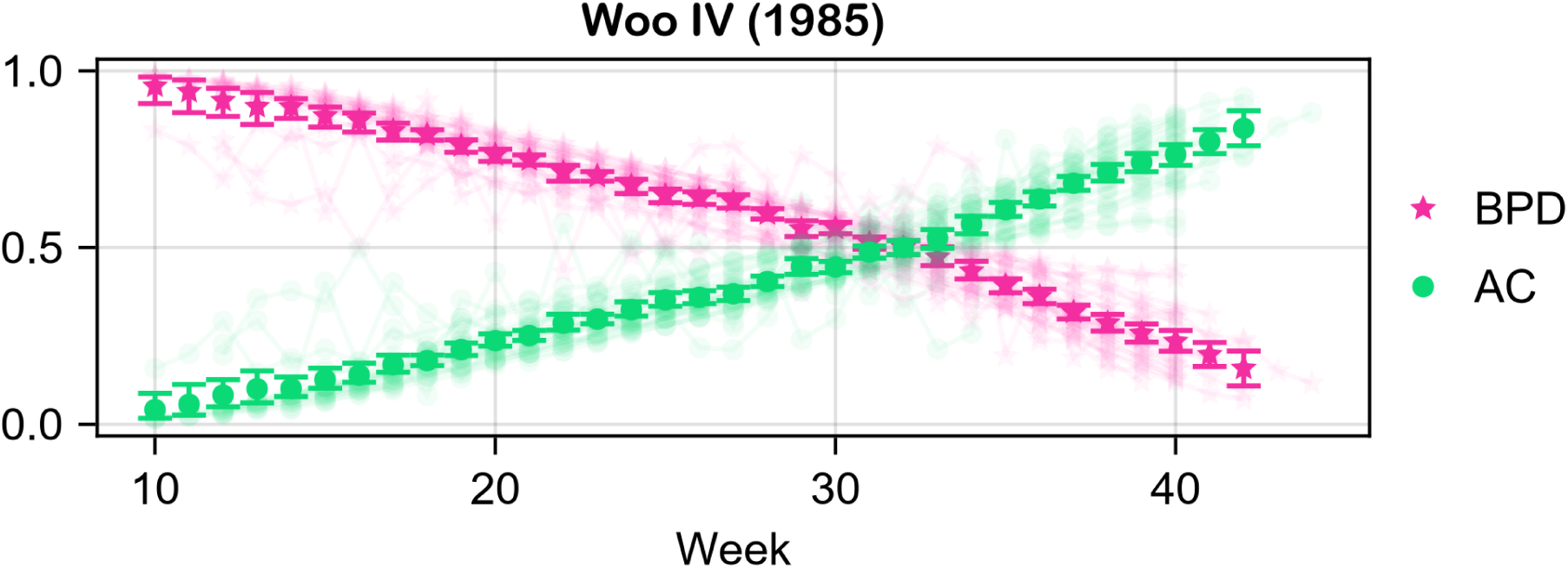
Estimate of the mean of the first-order Sobol’ indices of Woo IV (1985).

**Figure 31:**
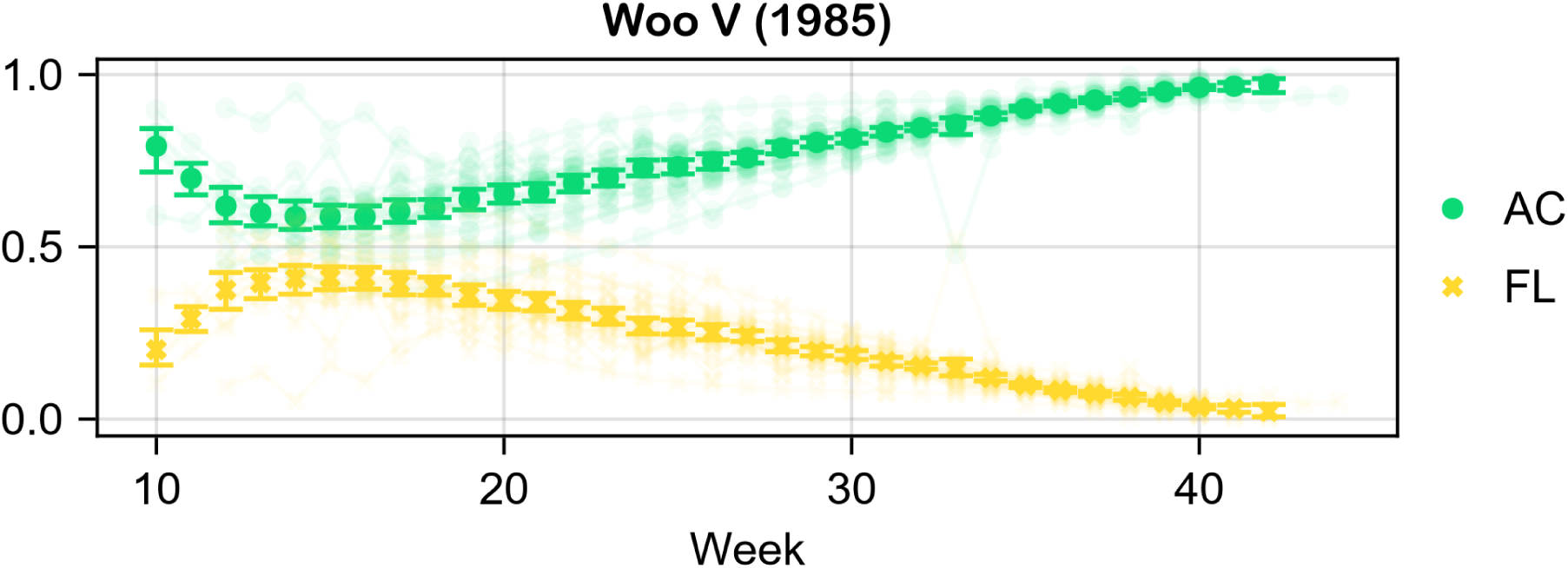
Estimate of the mean of the first-order Sobol’ indices of Woo V (1985).

The second-order Sobol’ indices for all the tested formulas were found to be small (*<* 0.01). Hence, they are not presented.

## 4 Discussion

### 4.1 Summary of results

In terms of a general overview of the results, approximately half of the formulas (45%) have at least one parameter with a first-order Sobol’ index that is less than 0.3. This indicates that half of the formulas are not parsimonious, as they utilize more parameters than are necessary to make a prediction for the EFW.

Moreover, 66% of the formulas exhibit a crossing of indices at some point during pregnancy. This indicates that in most formulas, the parameters undergo a reversal in their significance level.

With regard to the monotonicity of the indices, it can be observed that, following the 20th week, the vast majority of the studied formulas exhibit strictly monotone indices. During weeks 10 to 20, approximately 41% of the formulas exhibit at least one index with a turning point. Schild (2004) displays the turning point a couple of weeks after week 20. Notably, all of these formulas contain FL as a parameter. Furthermore, only two of the sixteen formulas with FL as a parameter do not display a turning point (12.5%).

With respect to the importance of each parameter, the presence of HC is insignificant (having an index less than 0.3 throughout the whole gestation) in 20% of the formulas that it appears in, BPD in 33%, AC in 4%, and FL in 38%. On the contrary, only 2 of the formulas (7%) have a dominant index, with values greater than 0.7 throughout the gestation. Specifically, HC is dominant in Weiner II (1985) and AC is dominant in Merz (1988). AC is generally considered to be a crucial parameter in fetal weight estimation. After all, it is included in all the studied formulas. The results of the present study highlight the importance of AC, however, just like all the tested parameters, its contribution depends on both the choice of the formula and the gestation age.

Regarding the second-order Sobol’ indices, they measure the degree of interaction between the associated parameters. The term “interaction” is used to describe a situation in which the effect of one variable on an outcome depends on the level of another variable. Consequently, if the second-order index between two parameters of a formula were to be identified as significant, it could, for instance, signify that one of the parameters must be above or below a specific threshold in order for the other to be taken into account. This scenario would be incongruent with the objective of fetal weight estimation, as the larger the value of a biometric parameter is, the larger the EFW should be, irrespective of the value of the remaining parameters. Hence, since none of the under investigation formulas admitted a significant second-order Sobol’ index, they are all well-defined.

As far as the uncertainty of the estimates is concerned, the length of the 95% CI is generally narrow, indicating a precise estimate. However, some formulas have a more broad 95% CI during weeks 10 to 15, such as INTERGROWTH-21 (2017) and Shepard I (1982), as well as during weeks 41 and 42, such as INTERGROWTH-21 (2017) and Halaska (2006). One potential explanation for the broader estimates is the smaller number of data points available for analysis in those weeks. As can be seen in Table 1, most of the datasets utilized do not provide data for the aforementioned weeks. This results in the bootstrap estimate being less reliable, thereby increasing the uncertainty, due to the smaller sample size. Another potential reason for this could be the variability of the data in those weeks. During early gestation, particularly between weeks 10 and 15, the fetus is still small, which can make accurate measurements of the parameters difficult^[44]^, thus, influencing the 10th and 90th percentile charts on which this study’s approach relies on.

### 4.2 Interpretations of results

To facilitate the interpretation of the results, some examples are in order. The following examples use data from Papageorghiou et al. ^[38]^, and concern week 38, which is an important week to predict fetal weight.

Assuming a fetus with the median biometric parameters, Hadlock III (1985) predicts the EFW to be 2990.85 g. When HC lies at the 90th percentile, with the remaining parameters lying at the median, then the EFW is 3102.99 g (3.7% increase). Doing the same for HC results in the EFW to be 3376.14 g (2.2% increase), whereas for AC the result is 3376.14 g (12.8% increase) and for FL is 3123.26 g (4.42% increase). The respective first-order Sobol’ indices for HC, AC and FL are 0.075, 0.82 and 0.103. It is obvious that the most substantial change occurs when AC is altered, followed by FL, and then HC.

Assuming a fetus with the median biometric parameters, Hadlock IV (1985) predicts the EFW to be 3049.05 g. When BPD lies at the 90th percentile, with the remaining parameters lying at the median, then the EFW is 3116.23 g (2.2% increase). Doing the same for HC results in the EFW to be 3116.92 g (2.2% increase), whereas for AC the result is 3444.67 g (12.97% increase) and for FL is 3173.29 g (4.07% increase). The respective first-order Sobol’ indices for BPD, HC, AC and FL are 0.027, 0.027, 0.853 and 0.09. It is evident that the most significant change occurs when AC is altered, followed by FL, and then BPD and HC, which both affect EFW similarly.

The results of the above practical examples demonstrate complete alignment with the findings of the present study.

### 4.3 Implications

The findings of the present study have immediate and significant clinical implications. For example, it can become increasingly challenging to accurately measure the parameters of the fetus’s head toward the later stages of pregnancy. Various factors, including the position of the fetus, the volume of amniotic fluid, and the presence of an anterior placenta, can influence the precision of these measurements. Furthermore, the fetal head may engage with the pelvis, potentially impeding the acquisition of clear ultrasound images^[34;60;41]^. Therefore, to minimize the error in predictions, it is recommended to use a formula with small first-order indices for HC and BPD.

### 4.4 Strengths and limitations

A notable strength of this study is its minimal reliance on a single, specific dataset of measurements, which could potentially be biased towards particular values. On the contrary, it relies solely on the estimates of the 10th and 90th percentile charts, allowing it to investigate the entire parameter space evenly, ensuring that the results are objective and impartial. Furthermore, the results are based on a bootstrap estimate of numerous datasets, with participants of various ethnicities, which provides additional assurance regarding the reliability of the findings.

A limitation of this study is the exclusion of outlier measurements from the explored parameter range. The omission of measurements scoring below the 10th and above the 90th percentile ensures the validity of the results for appropriate for gestational age fetuses. However, it should be noted that the respective indices for small and large for gestational age fetuses may vary from those presented in the current article. Hence, the dependence of fetal weight on individual biometric parameters for small and large for gestational age fetuses could be explored in a future study.

## 5 Conclusions

The above analysis leads to the following two conclusions:

1. The behavior of certain parameters of some formulas is subject to qualitative variation during pregnancy, as the values of the corresponding indices change dramatically. Therefore, each sonographer should take into account the gestational age in relation to the formula used for the fetal weight assessment.
2. Certain parameters of some formulas are insignificant throughout pregnancy, which has been confirmed in previous statistical studies^[46]^, such as the HC of Hadlock III (1985). Since fewer parameters are generally preferred (especially in a crude estimation on an emergency basis), some commonly used formulas should be reassessed in this context.

## Data Availability

The data and code that support the findings of this study are openly available on GitHub at github.com/TsilidisV/DopEFW.jl.

## Author Contributions

All authors contributed equally to this work.

